# Robotic-Assisted Gait for lower-limb Rehabilitation: Evidence of Altered Neural Mechanisms in Stroke

**DOI:** 10.1101/2022.02.01.22269218

**Authors:** Juan Manuel Mayor-Torres, Ben O’Callaghan, Attila Korik, Alessandra Del Felice, Damien Coyle, Sean Murphy, Olive Lennon

## Abstract

Robotic-Assisted Gait training (RAGT) offers an innovative therapeutic option for restoration of functional gait in stroke survivors, complementing existing physical rehabilitation strategies. However, there is a limited understanding of the neurophysiological response induced by this training in end-users. Neural desynchronization and Cortico-Muscular Coherence (CMC) are two biomarkers that define the level of muscle-cortex association during gait phases and can be used to estimate induced user’s adaptation during RAGT. In this study, we measure Event-Related Spectral Perturbation (ERSP) and CMC from three healthy individuals and three stroke survivors during overground-gait with and without an exoskeleton. Results show that (1) the use of the exoskeleton in healthy individuals is associated with a different and more refined motor-control represented in a high *θ*-desynchronization, (2) altered and noisy ERSP and lower and non-focal *β*-CMC patterns are observed in Stroke patients when performing overground-gait both with and without the Exoskeleton, and (3) Exoskeleton use in stroke survivors is associated with a reduction in swing-time during gait-cycle, but this effect is not correlated with an increment of *θ*-desynchronization and/or *β*-CMC. ERSP and CMC demonstrated evidence of neural modulation in able-bodied users during RAGT, which could not be detected in subacute stroke survivors during RAGT. These results suggest that the gait-parameters changes observed during exoskeleton use in subacute stroke survivors are unlikely to be neurally driven.

## I. Introduction

**S**TROKE is the second most common cause of death and a primary cause of adult acquired physical disability world-wide [1], [2], [3]. The Action Plan for Stroke Rehabilitation in Europe [4] stresses the need for high-intensity exercises for activities of daily living after stroke. Robotic-Assisted Gait Training (RAGT) [5], [3], [6], [7] has the potential to deliver intensive and progressive gait training in this population [8].

RAGT protocols have been proposed as an alternative to repetitive and isometric therapy. RAGT delivered during the early subacute phase after stroke [9] increases the likelihood of regaining independent walking and improves walking velocity and capacity. Exoskeleton devices further reduce the therapist’s involvement and the manual handling burden during active gait rehabilitation [10], [11], [12]. However, despite positive evidence supporting RAGT after stroke, it is not clear if RAGT devices provide a positive stimulation to promote neuroplasticity for restoration of independent and efficient walking [13], [14].

Electroencephalography (EEG) and Electromyography (EMG) recordings are informative, non-invasive measures of the activation levels of the neuromuscular system involved in human locomotion. Event-Related Spectral Perturbation (ERSP) [15], [16], [17] and Cortico-Muscular Coherence (CMC) [18], [19], [20] are EEG/EMG derived measures of neuromuscular plasticity. CMC is considered as a meaningful biomarker of the interactions between the motor cortex and the muscles involved in movement execution. Lower limb stroke rehabilitation studies investigating these biomarkers are lacking. Here we address this limitation by comparing ERSP and CMC biomarkers during gait, with and without the assistance of a RAGT device.

The majority of identified studies in stroke rehabilitation that report ERSP and/or CMC evaluate the activation of the upper limbs. Studies exploring lower limb activation typically describe healthy subjects, identifying a clear-cut synchronization-desynchronization (i.e., neural sync-desync) pattern during gait cycles [21], [22] as well as a high and contra-lateral CMC - observed more in the opposite hemisphere of the limb movement [23], [24]. These patterns are aligned with heel-strike (HS) and toe-off (TO) gait events. These studies identify different neural processing in healthy subjects during RAGT when compared to overground gait [22], [25]. Specifically, Knaepen et al. [25] reports that the increment of the percentage of allowed robotic guidance-force (GF) provided by an exoskeleton orthosis is associated with high neural-desynchronization between 2-45Hz during treadmill walking. A higher neural desynchronization in healthy individuals suggests a different and more refined motor-control that assists walking, compensating for new changes in the environment - in this case, the additional GF itself [26], [27]. Neural sync-desync (ERSP) patterns during overground gait described in individuals following stroke are altered in comparison with healthy individuals. Garcia-Cossio et al. [22] and Vinoj et al. [28] evaluated two different exoskeleton-assistance modes for lower-limb rehabilitation in healthy and stroke individuals. ERSP was not visually identifiable in stroke survivors during overground gait. Lower *θ, µ, β*, and *γ* sync-desync values were measured in stroke participants in comparison with the neural sync-desync pattern observed in healthy controls [25]. This finding suggests functional limitations after stroke in motor control of physiological walking [23], [24].

In CMC, stroke survivors show lower *β* and *γ*-CMC in comparison with healthy individuals [18], [29], [30], with an altered/poorer motor performance of affected limb signaled by lower *µ* and *β*-CMC values [31], [32], [33]. Bao et al. [32] reported reduced bilateral CMC from the Tibialis-Anterior (TA) muscle in stroke survivors in comparison with healthy participants. This CMC decrement was associated with the activation of source generators in the pre-central and cingulate cortices. Maggio et al. [33] reported CMC in stroke during RAGT identifying an increment of *β*-CMC from the TA in stroke survivors with lower-limb impairment after using a robot-aided ankle device - although the effect emerged only in robot-näive participants. No correlation was observed between the increment of *β*-CMC and improvements in gait-parameters.

Taken together, these findings suggest an association between higher neural sync-desync and lower limb muscle activation during gait in healthy participants [22], [28] but not in individuals following stroke when performing treadmill-walking [21], [34]. These findings have not been replicated in studies evaluating over-ground walking. While two studies were identified in stroke with co-registered EMG and EEG data during RAGT in a recent systematic review [5], it remains unknown if physiological neural sync-desync during gait can be regained through the support of RAGT after stroke. It has been proposed that evaluating the brain-muscle connection during active RAGT can allow estimation of the level of Exoskeleton/RAGT induced adaptability on and the potential utility for repetitive RAGT in the restoration of a functional and reciprocal gait pattern in individuals following stroke [3], [14].

In this study we evaluate the correlation between ERSP, CMC, and the phases of the gait-cycle in healthy individuals and stroke participants performing overground-gait (i.e. without treadmill use) with and without the use of a RAGT device. To aid our investigation of the adaptability of neural biosignals during RAGT and their potential to help restore a physiological gait pattern post stroke, our research questions were: 1) What are the gait-parameters, ERSP and CMC changes when healthy controls perform overground-gait (OG) with or without a RAGT device/Exoskeleton? 2) What are the corresponding gait-parameters, ERSP and CMC changes when stroke survivors perform OG and RAGT, and 3) Are the gait-parameters, ERSP, and/or CMC improving efficiently during RAGT as a tracking biomarker of recovery of a physiological gait pattern? i.e., is an observed change in gait-parameters during RAGT associated with increased neural activity as measured by ERSP and CMC?

This paper is structured as follows: (1) Materials and methods section explaining the experimental setup, signal acquisition, and ERSP and CMC definitions, (2) Results section presenting the ERSP and CMC plots, their corresponding statistical evaluation and correlations, (3) Discussion, and (4) Conclusions.

## II. Materials and Methods

### Experimental Setup

Participants (healthy controls and individuals in the early subacute phase of stroke) were recruited at the Mater Misericordiae University Hospital, Dublin, Ireland, as part of the EU-funded PROGAIT project. Institutional ethical approval was received prior to recruitment and written informed consent was received from participants. Six participants were included, three healthy controls and three individuals post-stroke. The corresponding demographics, lesion, and dominant hand are reported in Table I.

**TABLE I:**
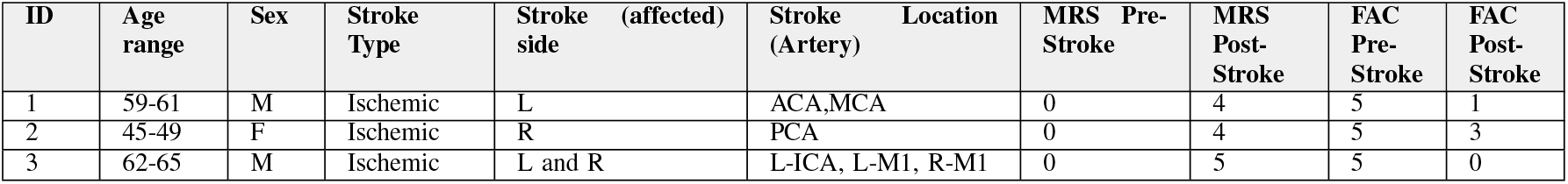
Stroke patients demographics and clinical information, including the Stroke (affected) side or the lesion side. The abbreviations in this Table are Modified Rankin Scale (MRS) [43], Functional Ambulatory Category (FAC) [44], Anterior Cerebral Artery (ACA), Middle Cerebral Artery (MCA), Left-Internal Carotid Artery (L-ICA), Myocardial Infarction (MI). MRS and FAC were scored the same day of the visit. All the stroke individuals were considered and screened under acute and/or subacute stage

Overground-gait trials were first conducted along a straight 20-meter walkway without the exoskeleton. For stroke survivors, these trials were conducted wherever feasible and to the best of the participants’ abilities. For robotic assisted gait, the trials were repeated using an Ekso GT(tm) overground Exoskeleton, Ekso Bionics, Richmond, CA, USA [35]. Participants were familiarized with donning and walking in the Ekso device in days prior to data collection. Participants’ anthropometric data dictated the Ekso dimensions which were individually adjusted by an Ekso-certified physiotherapist. The device was programmed to Adaptive-Assist Mode (AAM) before RAGT trials.

In this mode the actuators/motors provide assistance to the hip and knee joints only when a deviation from a programmed spatial trajectory is detected. RAGT trials were conducted on the same day with a rest period of up to fifteen minutes provided between walks.

### B. EEG, EMG, and Accelerometer recordings

The EEG signal was recorded in adherence with best EEG data-collection guidelines [36], [37], [38], [39], [40], [41] during overground walking using a wireless 32-channel gtec g.Nautilus active device, with a sampling rate of 250Hz. The EEG channels included were FP1, FP2, AF3, AF4, F7, F3, Fz, F4, F8, FC5, FC1, FC2, FC6, T7, C3, Cz, C4, T8, CP5, CP1, CP2, CP6, P7, P3, Pz, P4, P8, PO7, PO3, PO4, PO8, and Oz. Each EEG trial was referenced to the right-earlobe electrode and filtered using a band-pass Butterworth filter between 0.1-100Hz. The ground electrode was positioned over the AFz location according to the 10/20 EEG positioning.

Bipolar surface EMG and accelerometry data were acquired using the Delsys Trigno® device, Natick, MA, USA. EMG signals were sampled at 1925.93Hz and the accelerometer at 148.14Hz. EMG signals were obtained bilaterally from the Tibialis Anterior (TA), Soleus (SO), Rectus Femoris (RF), and Semitendinosus (St) muscles. All EMG sensors were positioned following the European Standards for Surface EMG sensors positioning (SENIAM) [42]. EMG signals were filtered using a Butterworth band-pass filter between 5-100Hz. This filtering was performed before the CMC calculation.

### C. EEG and EMG pre-processing

Motion and blinking artifacts were first removed from the filtered EEG signals using the Koethe’s cleanraw function from the Prep pipeline - specifically the Artifact Subspace Removal (ASR) [45]. The ADJUST plugin [46] from EEGlab [47] was then used to infer and remove artifactual independent components for each EEG trial using the pre-calculated Independent Component Analysis (ICA) decomposition matrix. ADJUST used Kurtosis and Skewness spatial thresholding to predict artifactual ICs and to subsequently remove them using an ICA composition. Canonical Component Analysis (CCA) was used to remove muscular artifacts [48]. For measuring how well this sequence of methods removed motion artifacts, we measured the Total Harmonic Distortion (THD) in *δ* (0.1-4 Hz) and *θ* (4-7Hz) rhythms finding significant differences F(1,)=13.35, p=0.0278 - always obtaining a lower THD after applying this artifact removal [49]. For checking the results of the THD analyses refer to the supplementary material.

### D. Event-Related Spectral Perturbation (ERSP)

Event-Related Synchronization (ERS) and Event-Related Desynchronization (ERD) measures represent the localized increase and decrease in the amplitude of rhythmic activities in association with one or multiple stimuli/events [52], in this case a gait-cycle event and a baseline spectrum related to an EEG baseline period. This baseline spectrum was calculated from the EEG signal contained within 0.5 seconds before each gait-event [16], [17]. In gait analysis, the ERSP was calculated to identify the level of continuity of motor-control associated with Left and Right HS and TO events, indicating transitions to stance and swing phases [17], [53].

For calculating ERSP, we defined an EEG spectral estimate as *F*_*k*_(*f, t*) where *f* denotes frequency, *t* time and *k* the index of a trial belonging to a particular gait event. To analyze ERSP we defined gait events in four different phases, e.g., LTO-LHS (i.e., the left swing phase), LHS-RTO (Left double limb stance) or RTO-RHS (right swing phase), RHS-LTO (right double limb stance). LTO-LTO or RTO-RTO phases represent the 100% of the gait-cycle i.e. left or right full stride. The HS event refers to the beginning of the stance phase on each limb. To generalize the spectral perturbation, the additive ERSP model was introduced [54] in line with similar multiple EEG analyses [3], [6]. Equation 1 defines ERSP in terms of the time on the EEG baseline excerpt *t*′, and the spectral perturbation as an average normalization of *F*_*k*_(*f, t*) per trial, and the mean and standard deviation of the baseline spectrum *F*_*k*_(*f, t*′), denoted as *µ*_*B*_ and *σ*_*B*_.

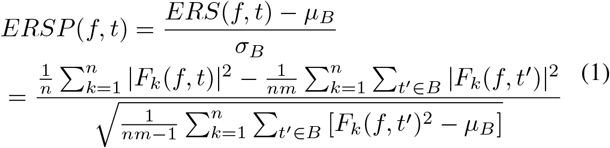

When measuring ERSP at signal level, ERSP was transformed from normalized values in percentages to decibels (dB) with a purpose of standardized ERSP measurement [54]. A positive ERSP was deemed to represent a synchronization or a matching between *F*_*k*_(*f, t*) and *F*_*k*_(*f, t*′), otherwise, it represented a desynchronization or a mismatch between *F*_*k*_(*f, t*) and *F*_*k*_(*f, t*′). In this study, we focused more on desynchronization spots which are directly related to motor-control transitions during normal gait [23], [17].

The ERSP was calculated using the *newtimef* function in EEGlab. We set a 0.8 s Hanning window [3], [17], [15] in the time-domain for the ERSP segmentation. An overlap or pad-ratio of 0.13s was used in each ERSP calculation to minimize discontinuities in the ERSP calculation.

The heel-strike (HS) and toe-off (TO) events, signaling the beginning of stance and swing phases, were identified using the amplitude local maxima from the shank accelerometer based on the Teager Kaiser Energy Operator (TKEO) [50]. To remove data periods where subjects were not actively walking, we used an automatic bouts-detection algorithm based on the rectified Signal Vector Magnitude (SVM) representation from the shank accelerometer. The rectified SVM located bouts indexes using the adaptive Otsu’s threshold - typically used for smoothing image edges [51].

### E. Cortico-Muscular Coherence (CMC)

Cortico-Muscular Coherence (CMC), a measure of the functional connection between the motor cortex and the muscle, was next considered [29]. In previous studies [54], [55], the correlation between two physiological time-series has been associated with the neural spiking-coupling between a neural process *N* and a peripheral signal *x*. CMC can be defined as a spectrum-based approximation of the Granger causality between an EEG time-series associated with the neural process *N*, denoted as *y*, and an EMG signal *x* [55].

*Defining the auto-spectrum of x* as *f*_*xx*_(*ω*), and the cross-spectrum between *x* and *y* as *f*_*xy*_(*ω*) - with *ω* as the frequency domain, a bounded and normalized measure for the association between *N* and *x*, denoted as *R*_*xy*_(*ω*) can be established in Equation 2.

*R*_*xy*_(*ω*) is the CMC between the EEG time-series *y* and the EMG signal. This measure is bounded between 0 and 1, 1 being the maximum coupling between *x* and *y*, and 0 no ║ coupling. operator is an absolute value, and *·* operator is a dot-product between *f*_*xx*_(*ω*) and *f*_*yy*_(*ω*).

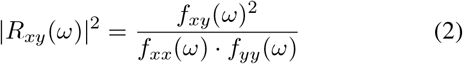

For obtaining the cross and auto spectrums, we used the *xspectrogram* function in Matlab 2020a. We used the psd flag/option, and a windowing of 0.5s with an overlap of 0.04s [30]. CMC was then obtained for each gait phase, and between each EEG channel and the muscles

### F. Data Analysis

Data are reported separately for healthy controls (HC) and stroke participants (ST). For stroke participants, we refer to the stroke side as the lesioned hemisphere, i.e. the left stroke side refers to right limb impairment, and the right stroke side refers to left limb impairment. Differences between the walking conditions (overground gait with and without exoskeleton assistance) were evaluated using one-way ANOVA tests, with F and p-values reported between the walking conditions for gait-parameters, ERSP and CMC values. For simplicity, some significant differences were reported using F and p-values as intervals. All p-values reported in pairwise comparisons were adjusted using the Bonferroni-Holm correction.

For ERSP analyses, pairwise comparisons refer to LTO-LHS (left swing time), LHS-RTO (Left stance time), RTO-RHS (right swing time), and RHS-LTO (right stance time) gait-events, with an assumption that the ERSP values are associated with these gait-events and not to a particular limb [21], [22]. For CMC statistical comparisons, CMC values within a 95% average confidence interval on each frequency band were pre-selected [30], [31]. The CMC axis limits were then re-defined between [0.12,0.15] as we specify in Figure 2a. A Generalized Linear Model (GLM) was used for ERSP analysis and we report a simple intercept and slope estimation.

## III. Results

Results for stroke participants are reported for the unaffected or stroke/affected side where a significant difference appears on that side.

### A. Participant Demographics

Three healthy participants (two female; aged 36.0 ± 12.12 years), and three individuals (one female; aged 57.0±8.71 years) in the early subacute phase after an ischemic stroke were included. One individual post-stroke was unable to walk without the Exoskeleton assistance. No EEG or EMG data were registered for OG walking in this participant. Table I presents a summary of stroke participant demographics and clinical profiles including the Modified Rankin Scale (MRS) for disability and the Functional Ambulatory Category (FAC) [43], [44]. See Table I for more details about ST demographic and clinical measures.

### B. Comparison of-Overground-Gait v.s RAGT in Healthy Controls (HC) and individuals after Stroke (ST)

#### 1) Gait-parameters

**HC:** Significant differences were identified in gait parameters and in healthy individuals between OG and exoskeleton-assisted trials in stride times (LTO-LTO), F(1,255)=80.12, p=4.144E-6, in swing-time, e.g. LTO-LHS, F(1,281)=96.55, p=8.995E-8, and RTO-RHS, F(1,281)=55.89, p=6.442E-5, and stance-time but only on the right side RHS-LTO, F(1,281)=21.57, p=0.00045. Times were shorter in exoskeleton gait in comparison with OG trials only in swing and stride and the right stroke side. Table II provides a detailed breakdown by gait phases during the two modalities OG and Exoskeleton assisted gait (Ekso). Healthy individuals walked a mean of 28 walking trial bouts during overground gait, and an average of 5 bouts using the exoskeleton.

**TABLE II:** Average and standard-deviation describing gait-performance in times - in seconds. The times reported here are associated with healthy controls (HC) or Stroke Patients (ST) performing Overground-Gait (OG) or Ekso exoskeleton-assisted trials. The left or right stroke side is referring to the lesion side in the motor-cortex and the clinical information about the lesion is reported in Table 1. The opposite lower limb is impaired in relation with the lesion side. * represent a significant improvement, F(1,201) ≥ 12.77, p ≤ 0.00389, in the gait performance associated with Stroke patients between the OG and Ekso modalities.

| Time [s] |  | Left Stroke side (Right Affected Limb) |  |  |  |  | Right Stroke side (Left Affected Limb) |  |  |  |  |
| --- | --- | --- | --- | --- | --- | --- | --- | --- | --- | --- | --- |
|  |  | LTO-LTO* | LHS-RTO | LTO-LHS | RHS-RTO | RTO-RHS* | LTO-LTO* | LHS-RTO | LTO-LHS* | RHS-RTO | RTO-RHS |
| OG | HC | 1.247±0.193 | 1.543±3.912 | 0.545±0.254 | 0.226±0.665 | 0.593±0.225 | 1.152±0.531 | 0.205±0.613 | 0.532±0.203 | 1.513±4.112 | 0.516±0.246 |
|  | ST | 5.442±2.887 | 1.485±3.447 | 0.777±0.284 | 2.485±1.557 | 1.132±0.663 | 3.225±0.531 | 1.334±0.514 | 0.586±0.187 | 0.986±0.252 | 0.545±0.367 |
| Ekso | HC | 3.824±1.112 | 1.775±4.112 | 0.846±0.445 | 1.487±1.007 | 0.723±0.275 | 3.951±0.987 | 1.485±0.778 | 0.712±0.336 | 1.886±3.131 | 0.768±0.266 |
|  | ST | 3.278±1.099 | 1.113±0.924 | 0.789±0.751 | 2.887±3.221 | 0.604±0.254 | 2.203±1.107 | 2.525±3.552 | 0.517±0.487 | 1.223±0.485 | 0.522±0.244 |

**ST:** Significant differences in stride times (LTO-LTO) between OG and Ekso modalities on the stroke side, consistent for Left stroke side F(1,220)=98.84, p=8.823E-9, and Right stroke side, F(3,220)=22.73, p=0.00039, emerged, with Ekso times consistently shorter than OG. Significant differences between OG and Ekso modalities during swing-time for the stroke affected limb were also evident in RTO-RHS F(3,220)=15.64, p=0.00358, and in LTO-LHS F(3,220)=16.92, p=0.00178, with Ekso swing-times shorter than OG.

Non-significant differences were observed during swing times in the unaffected limbs; (RTO-RHS) F(3,220)=9.88, p=0.0867, and (LTO-LHS) F(3,220)=8.56, p=0.1011. See Table II for details. Stroke individuals walked a mean of 4 trial bouts during overground gait, and an average of 4 bouts using the exoskeleton device.

*ERSP:* ERSP measures were grouped according to gait cycle phase (LTO-LHS, LHS-RTO, LTO-LTO for the left, and RTO-RHS, and RHS-LTO and RTO-RTO for the right limb). **HC:** Pairwise comparison of overground versus exoskeleton assisted gait identified significant differences in *γ* (30-50Hz) desynchronization during stance-time in Cz,F(1,5) ≥ 19.39, p ≤ 0.0386, with OG<Ekso. This disparity in findings means that the ERSP values in *γ* were significantly lower or more negative in OG than Ekso trials, i.e. the *γ* desynchronization observed during OG was significantly higher than Ekso. Differences in *γ* were further observed in C3, C4 and Pz; F(1,5) ≥16.78, p≤ 0.0337; F(1,5) ≥14.88, p≤ 0.0395, and F(1,5) ≥30.578, p ≤0.0167, respectively, again all OG<Ekso. Significant differences were also observed in *θ* (4-7Hz), during the stance phase at Cz F(3,5) ≥23.22, p≤ 0.0256, C3 F(3,5) ≥13.98, p≤ 0.0401, C4 F(3,5) ≥ 17.63, p ≤0.0302, and Pz F(3,5) ≥37.38, p ≤ 0.007. However, in these instances all comparisons showed OG*>*Ekso meaning *θ*-desynchronization observed in Ekso trials is significantly higher than in OG trials. This higher *θ*-desynchronization is directly associated with the usage of the Exoskeleton in HC.

No significant differences were observed in other frequencies or during the swing-phases of gait F(1,5) ≤ 3.323, p*>*0.05. **ST:** In contrast to the findings reported in healthy controls (HC), for stroke participants no desynchronization was apparent during gait events as shown in the middle and bottom ERSP plot in Figure 1. No significant differences were observed for any frequency or during any gait phase, F(1,5) ≤ 3.769, p*>*0.05 when comparing OG and Ekso modalities in stroke participants.

**Fig. 1:**
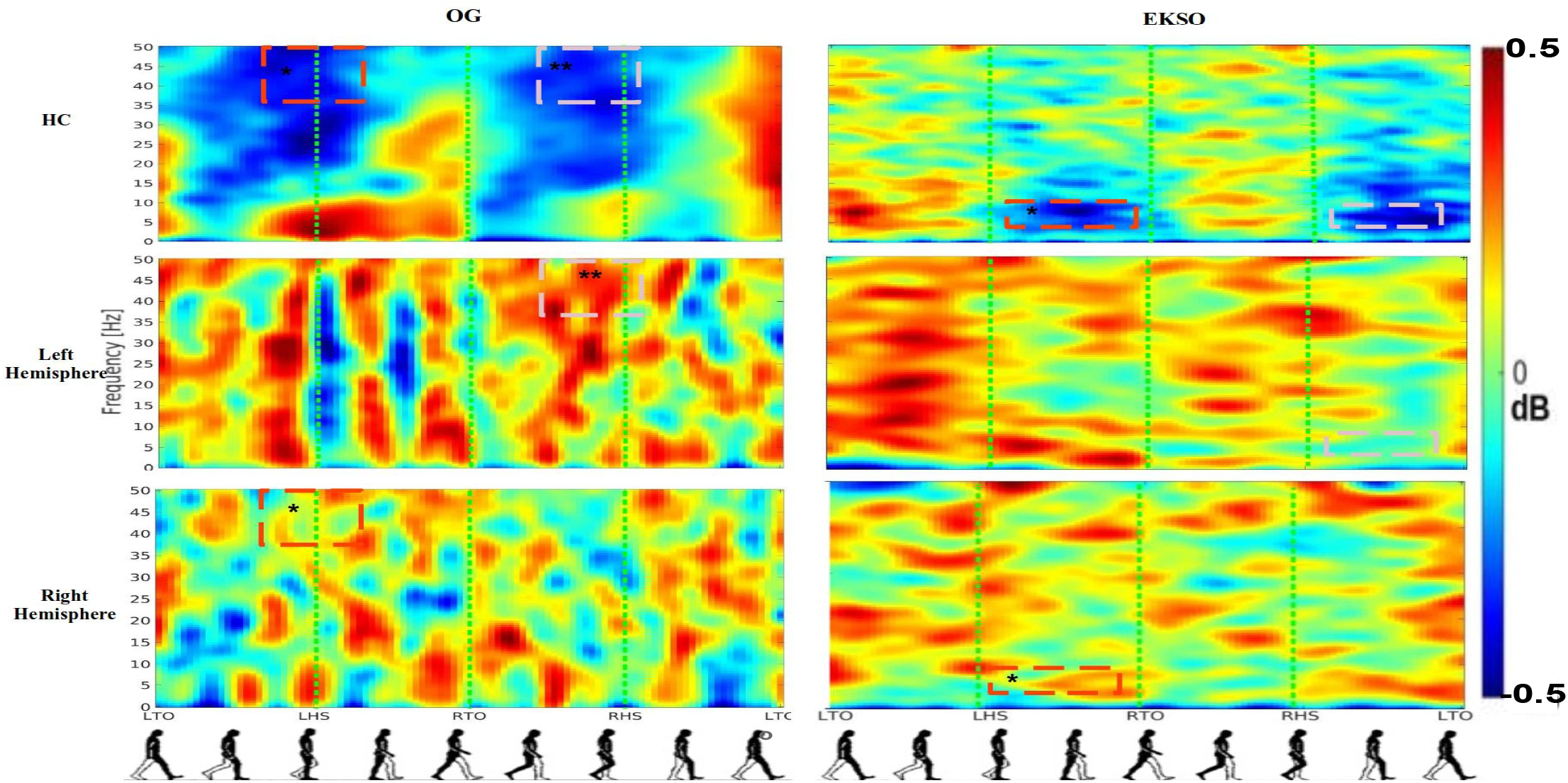
Average ERSP output for the Cz, C3, C4 channels. This Figure shows in rows the groups - healthy-controls (HC), and Stroke Patients (ST) Left Stroke side and Right Stroke side, and in columns the gait-evaluation modalities OG and Ekso. Each plot has in y-axis frequency between 0-50Hz, and in x-axis all the gait-phases of the gait-cycle. Red spots represent synchronization and darker blue spots represent desynchronization. The level of sync-desync is defined in the colorbar between [-0.5,0.5] dB and plotted based on the jet colormap. The red and pink dashed spots represent differences between the well-defined desynchronization spots found in HC and with the noisy sync-desync patterns found in ST. This has been observed before in Garcia-Cossio et al. [22]. These differences are more pronounced in the affected limb. Comparing OG and Ekso modalities we can see an evident difference in the desynchronization values - being allocated in the *γ* rhythm for OG, and in the *θ* and *µ* rhythms for Ekso. This is also suggesting a different on the neural processing responsible for controlling walking task after exoskeleton use as Knaepen et al. [25] suggested. Red and pink spots represent a comparison between HC and ST groups on the same walking condition i.e., OG or Ekso.

**Fig. 2:**
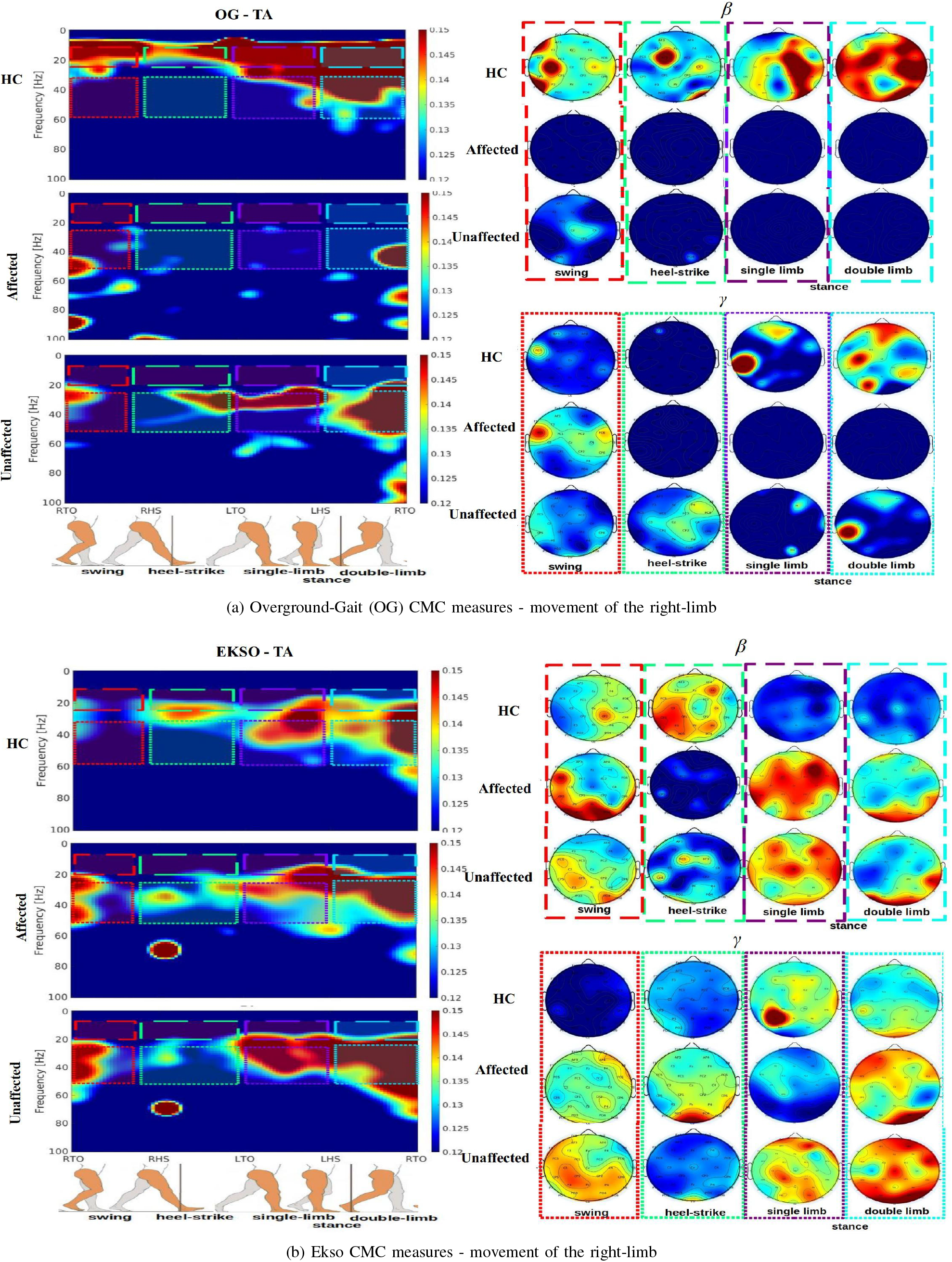
Average CMC measures for Left Stroke Side, reporting the CMC output for healthy-controls (HC), and Stroke patients (ST) for Affected and Unaffected limb. This output is only associated with the movement of right lower-limb. All the CMC values are plotted between [0.12,0.15] using the jet colormap and respecting the 95% of confidence interval described in we followed the methdology in [30], [31]. Figures 2a and 2b show the CMC outputs for the OG and Ekso modalities. In left plots we reported the CMC setting y-axis as frequency in Hz and x-axis as the gait-phases associated with the movement of the right limb. The gait-phases reported here are the swing, the heel-strike (HS), including the values in RHS-LTO, and the full stance time LTO-RTO. The topoplots showed in the right panel represent the average CMC values for swing, heel-strike, and single-limb stance, and double-limb stance intervals for *β* and *γ* rhythms. A more focal posterior activity is observed when healthy-controls walk with the exoskeleton (EKSO) or analyzing the movement of the unaffected limb for stroke patients in comparison wih OG modalities with CMC activity focuses on contra-lateral (e.g., opposite to the moving limb) central regions.

#### 3) CMC

CMC results are grouped as four different gait-phases relative to the motion of a limb: *swing* (LTO-LHS, RTO-RHS), *heel-strike* (LHS-RTO, RHS-LTO), *single-limb stance*, when the opposite limb is non-weight bearing during swing, (LTO-LHS, RTO-RHS), and *double-limb stance*, a short phase when the opposite limb is performing heel-strike (LHS-RTO, RHS-LTO). Figure 2 shows this gait cycle segmentation graphically.

**HC:** The first row/panel of Figure 2a and 2b show *β* (16-30Hz) and *γ*-CMC measured from the TA muscle for OG and Ekso modalities, respectively. For HC we observed significant differences in right-limb swing-time for *β*-CMC and C3, F(1,5) ≥18.325, p≤ 0.0225, and in left-limb swing-time and C4, F(1,5) ≥19.886, p ≤ 0.0192. In these comparisons we measured higher CMC values in OG in comparison with Ekso (OG*>*Ekso).

The heel-strike (HS) period showed similar significant differences in *β*-CMC and C3, for the right-limb, F(1,5) ≥ 20.734 p≤ 0.0106, and C4 for the left-limb, F(1,5) ≥ 22.561, p≤ 0.00882 in TA muscle. Comparisons showed higher CMC values in OG. Further differences were identified during single-limb stance time in *β*-CMC at C3, in the right, F(1,5) ≥ 11.771 p≤ 0.0327, and at C4 in the left single-limb stance, F(1,5) ≥ 10.446, p≤ 0.0349, again with higher CMC values in OG. Other differences were observed after grouping the *β*-CMC values in C3, Cz, and C4 channels and in HC. These differences are observed for the TA muscle F(1,5) ≥ 35.27, p ≤ 0.0125, with OG*>*Ekso and RF muscle F(1,5) ≥ 22.64, p≤ 0.0308 with OG<Ekso in the single-limb stance phase. No significant differences were observed during double-limb stance, *γ*-CMC, other channels or muscle group F(1,5) ≤ 2.881, p*>*0.05.

**ST:** During heel strike, significantly higher *β*-CMC for the TA muscle was observed in EKSO walking. These differences were observed contralaterally, at PO3 in participants with Left Stroke side F(1,5) ≥ 13.76, p≤ 0.0308, and at PO4 for those with a Right Stroke side F(1,5) ≥30.24, p ≤0.0112. Similarly, *β*-CMC was higher during EKSO walking in Pz from the TA F(1,5) ≥ 24.41, p≤0.0226, the SO F(1,5) ≥ 20.72, p≤ 0.0317, and the RF muscle F(1,5) ≤ 19.38, p≤ 0.0387 during the heel-strike and single-limb stance in the affected hemisphere - being OG<Ekso.

Overall a non-focal *β*-CMC was observed during RAGT. A lower but more focal OG *β*-CMC was observed in ST in comparison with HC. This effect is clearly evident in the first panel of Figure 2b [12], [56].

The maximum of *β*-CMC is observed in parietal-occipital regions when stroke patients perform walking with the EKSO in comparison with OG. This effect can be seen on panel two and three on Figure 2 this effect is more observable in TA and RF muscles - see supplementary material. No significant differences are observed in *γ*-CMC or other muscle groups during other gait phases, F(1,5) ≤ 4.751, p*>*0.05. More ERSP and CMC patterns for other channels, such as, C3, C4, and Pz, and other muscles, such as, RF, and SO are described in supplementary material.

### C. Correlation Analysis

The R and p-values reported in this section contain the effect contribution of both HC and ST groups separately, with only the noteworthy findings reported. Table III provides individual R and p-values of the correlations conducted comparing swing and stance times with the corresponding *β*-CMC values.

**TABLE III:** R and p-values for all the correlations between the gait-performances (times) - swing and stance phases and the *β*-CMC values on swing and stance phases per trial. Correlation results are reported for Unaffected and the Affected limb corresponding to the opposite limb in terms of the lesion side - in columns. In rows we reported the two different gait modalities such as OG and Ekso. SO shows negative correlation between the *β*-CMC values and the single-limb stance times when using the Ekso exoskeleton in both Unaffected and Affected limbs. RF shows a negative correlation with the stance times too, but only in the affected limb. No significant correlations are observed for St.

| Unaffected |  | TA Left ST side |  | TA Right ST side |  | SO Left ST side |  | SO Right ST side |  | RF Left ST side |  | RF Right ST side |  |
| --- | --- | --- | --- | --- | --- | --- | --- | --- | --- | --- | --- | --- | --- |
|  |  | swing | stance | swing | stance | swing | stance | swing | stance | swing | stance | swing | stance |
| OG | R | -0.058 | -0.173 | -0.028 | -0.035 | 0.169 | -0.025 | 0.103 | 0.003 | -0.138 | -0.016 | -0.141 | 0.096 |
|  | p | 0.487 | 0.041 | 0.762 | 0.676 | 0.064 | 0.552 | 0.275 | 0.965 | 0.101 | 0.844 | 0.106 | 0.253 |
| Ekso | R | -0.053 | -0.112 | 0.099 | **-0.164** | -0.133 | **-0.221** | 0.084 | **-0.287** | -0.059 | 0.041 | -0.028 | 0.051 |
|  | p | 0.518 | 0.179 | 0.181 | **0.039** | 0.081 | **0.007** | 0.301 | **0.0003** | 0.465 | 0.616 | 0.729 | 0.528 |
| Affected |  | TA Left ST side |  | TA Right ST side |  | SO Left ST side |  | SO Right ST side |  | RF Left ST side |  | RF Right ST side |  |
|  |  | swing | stance | swing | stance | swing | stance | swing | stance | swing | stance | swing | stance |
| OG | R | -0.047 | **0.191** | -0.086 | **0.259** | -0.022 | 0.033 | -0.123 | 0.077 | -0.128 | -0.065 | -0.142 | -0.128 |
|  | p | 0.615 | **0.034** | 0.306 | **0.005** | 0.813 | 0.726 | 0.07 | 0.411 | 0.171 | 0.492 | 0.078 | 0.139 |
| Ekso | R | 0.112 | 0.135 | -0.076 | -0.111 | 0.062 | **-0.275** | -0.093 | **-0.168** | 0.045 | **0.365** | 0.006 | **0.3** |
|  | p | 0.146 | 0.109 | 0.337 | 0.139 | 0.455 | **0.001** | 0.239 | **0.034** | 0.596 | **6.55e-6** | 0.937 | **0.00017** |

Significant negative correlations were observed only between the stance times (not swing times) reported in Table IV and the *β*-CMC grouping the values in C3, C4, and Cz for exoskeleton walking - always in the SO muscle in HC and in the unaffected and affected limb in ST. We also observed negative correlations between the stance-times and *β*-CMC from the TA muscle and the right stroke side but only in the unaffected limb. There was no significant correlation identified using ST measures only.

**TABLE IV:**
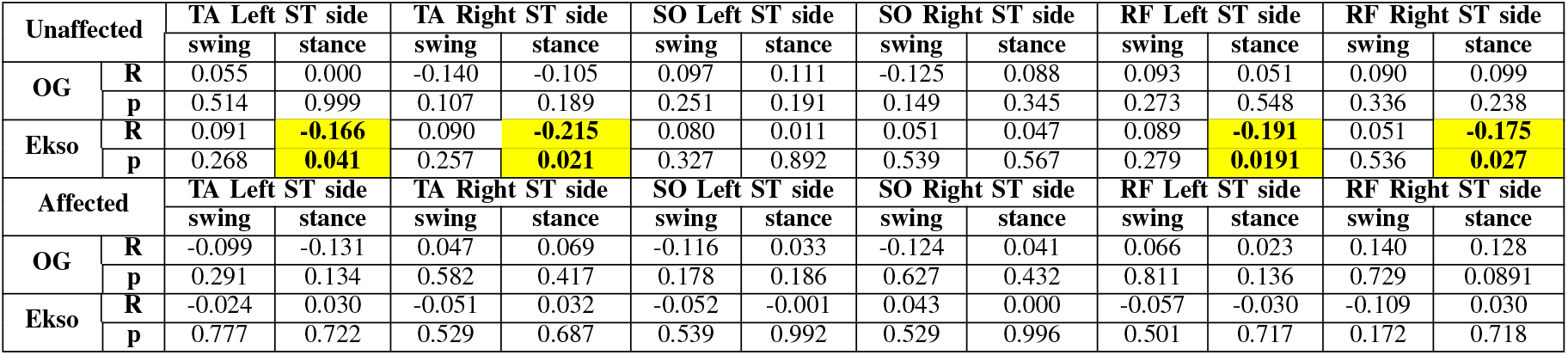
R and p-values for all the correlations between the *θ* and *γ* ERSP, for OG and Ekso, and the *β*-CMC values on swing and single-limb stance phases per trial and grouping the values on C3, C4, and Cz channels. Correlation results are reported for Unaffected and the Affected limb corresponding to the opposite limb in terms of the lesion side - in columns. In rows we reported the two different gait modalities such as OG and Ekso. TA and RF shows negative correlation between the *β*-CMC values and the *θ* ERSP using the Ekso exoskeleton, but only in the Unaffected limb. Values in bold show a significant negative correlation between *θ* ERSP and *β*-CMC grouped on C3, C4, and Cz. St measures are not reported here and none of them show significant correlations. No significant correlations are observed in the swing-time and/or for the other muscles.

| Unaffected |  | TA Left ST side |  | TA Right ST side |  | SO Left ST side |  | SO Right ST side |  | RF Left ST side |  | RF Right ST side |  |
| --- | --- | --- | --- | --- | --- | --- | --- | --- | --- | --- | --- | --- | --- |
|  |  | swing | stance | swing | stance | swing | stance | swing | stance | swing | stance | swing | stance |
| OG | R | 0.055 | 0.000 | -0.140 | -0.105 | 0.097 | 0.111 | -0.125 | 0.088 | 0.093 | 0.051 | 0.090 | 0.099 |
|  | p | 0.514 | 0.999 | 0.107 | 0.189 | 0.251 | 0.191 | 0.149 | 0.345 | 0.273 | 0.548 | 0.336 | 0.238 |
| Ekso | R | 0.091 | **-0.166** | 0.090 | **-0.215** | 0.080 | 0.011 | 0.051 | 0.047 | 0.089 | **-0.191** | 0.051 | **-0.175** |
|  | p | 0.268 | **0.041** | 0.257 | **0.021** | 0.327 | 0.892 | 0.539 | 0.567 | 0.279 | **0.0191** | 0.536 | **0.027** |
| Affected |  | TA Left ST side |  | TA Right ST side |  | SO Left ST side |  | SO Right ST side |  | RF Left ST side |  | RF Right ST side |  |
|  |  | swing | stance | swing | stance | swing | stance | swing | stance | swing | stance | swing | stance |
| OG | R | -0.099 | -0.131 | 0.047 | 0.069 | -0.116 | 0.033 | -0.124 | 0.041 | 0.066 | 0.023 | 0.140 | 0.128 |
|  | p | 0.291 | 0.134 | 0.582 | 0.417 | 0.178 | 0.186 | 0.627 | 0.432 | 0.811 | 0.136 | 0.729 | 0.0891 |
| Ekso | R | -0.024 | 0.030 | -0.051 | 0.032 | -0.052 | -0.001 | 0.043 | 0.000 | -0.057 | -0.030 | -0.109 | 0.030 |
|  | p | 0.777 | 0.722 | 0.529 | 0.687 | 0.539 | 0.992 | 0.529 | 0.996 | 0.501 | 0.717 | 0.172 | 0.718 |

No significant negative or positive correlations were observed between swing-time and *β*-CMC, regardless of group. No significant correlations were observed between any other gait-parameter and *γ*-CMC.

Table IV reports the R and p-values related to the GLM constructed between the *β*-CMC of C3, C4, and Cz and the *γ* and *θ* desynchronizations associated with the OG and Ekso modalities, respectively.

Significant negative correlations are observed in TA and RF muscles in the single-limb stance period but only from the unaffected limb in stroke. Evaluating the HC values only, this negative correlation is more significant R=-0.587, p=0.0067, and it is non-significant when analyzing the ST measures only (0.0135 ≤ R ≤ 0.0223, p*>*0.05).

Figure 3 shows the plots of the GLMs between *β*-CMC, in y-axis, and *θ* ERSP, in x-axis for Ekso walking. This negative correlation signifies a significant relationship between high *θ*- desynchronization values from central electrodes and high *β*- CMC obtained from TA and RF muscles of the unaffected limb. These correlations are strongly associated with the HC data where the negative correlation is preserved and increased as we observed above. No significant correlations were observed between *θ*-desynchronization and *β*-CMC in the SO and St muscles for HC and/or ST p*>*0.05.

**Fig. 3:**
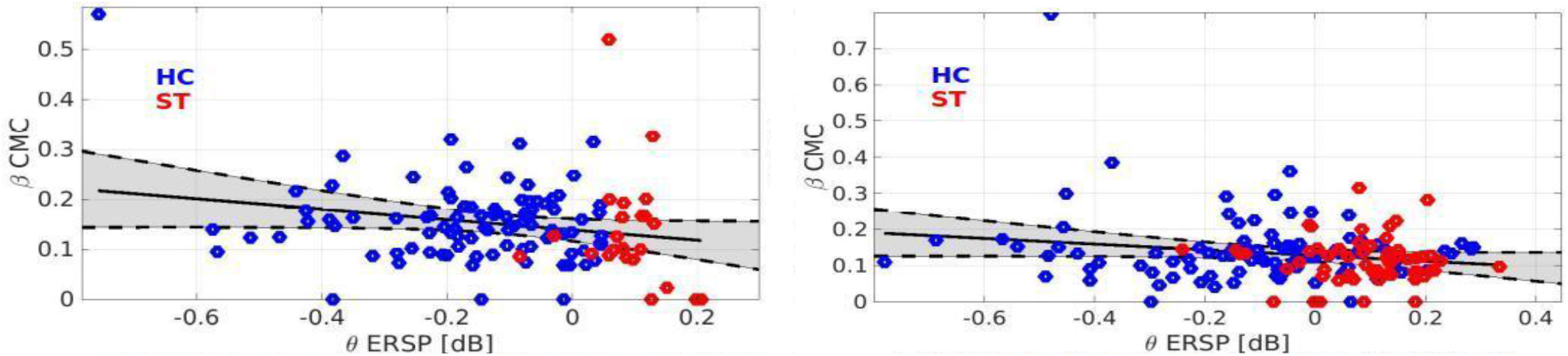
GLM analysis evaluating *β*-CMC, in the y-axis, and *θ* ERSP values associated with the inclusion of Ekso exoskeleton in the x-axis. X-axis is represented as normalized *θ* sync-desync values. R and p-vaues are reported in the captions correspondingly. HC points are plotted in blue, while ST points are plotted in red - all of them representing individual trials. Negative correlations are observed for TA and RF muscles, thus associating the *β*-CMC measured in the *single-limb stance* time with the high *θ*-desynchronization related to different motor control - but only in HC.

## IV. Discussion

The results reported in this study suggest that there are modifications of neuromuscular response induced by exoskeleton assisted gait modifications. Alterations of reduction in some gait-parameters, especially in the swing-time, observed in participants with stroke during exoskeleton use is not correlated with any increment of *θ*-desynchronization and/or *β*-CMC. Specifically, during the single-limb stance time, the brain-to-muscle connection of the affected lower limb appears less efficient.

Effectiveness of robotic gait training devices to drive neuroplastic recovery of physiological gait in stroke survivors is still unclear [5], [16]. While evidence supports the use of these devices to improve gait-performance compared with treadmill or free-walking training [11], [12], the potential to evoke adaptive neuroplastic changes has rarely been addressed [56], [57]. ERSP and CMC in healthy individuals during unassisted overground walking and exoskeleton assisted walking identified a neural sync-desync pattern aligned, respectively with Heel Strike and Toe Off. In Ekso walking, there is an evident and significantly higher desynchronization in *θ* rhythm in comparison with OG. These results are in line with previous findings using treadmill-based walking protocols [21], [22]. Frequencies involved in motor-control during healthy walking [24], [57] were recorded predominantly in *θ* and *γ* during walking with and without exoskeleton.

The increase of slower rhythms during exoskeleton walking suggests attentional/postural ongoing tasks, implying that the addition of the exoskeleton assistance elicits a different neural-processing during RAGT in healthy controls [33], [57]. The altered motor demands imposed by the use of the Ekso exoskeleton device, as well as the higher cognitive load imposed by a different step initiation dynamics compared to physiological gait, may explain the higher *θ* component during EKSO walking. This higher *θ*-desynchronization may be interpreted as an attentive midline *θ* [58], *which has been described during postural tasks [57], [58]. The activation of different scalp areas using the exoskeleton suggests a neural reorganization in healthy individuals, and reinforces the subsequent activation of related spinal-cord activations reflected in the CMC [34], [58]. High β*-CMC is observed in TA, SO and RF being more focal in HC and appears more clearly on central channels such as C3, Cz, and C4. This suggests a clear activation of the motor cortex during swing and stance phases [16], [19], [26], [59], and during single-limb and double-limb stance phases - see Figure 2.

Next, we investigated ERSP and CMC changes during free overground walking and EKSO walking in stroke survivors. We observed a different ERSP and CMC in ST compared with HC during unassisted and exoskeleton-assisted overground walking, as observed in [19], [56], [57]. The different neural activation patterns may be suggestive of the presence of ongoing neuroplasticity changes in M1, SMA and SII, which are responsible for physiological locomotion [56], [57].The neural desynchronization pattern is not clearly identifiable in post-stroke participants. This is consistent with previous studies, such as Garcia-Cossio et. al [22], where the neural desynchronization pattern observed in individuals with stroke was altered and non-aligned with any gait phase. In this study *β* and *γ*-CMC show some short and abnormal desynchronizations in stroke patients and in OG and EKSO modalities, but only during unaffected limb movements.

The altered sync-desync pattern observed in stroke survivors in this study is accompanied by a lower and sparse *β*-CMC.

During OG, the *β*-CMC is low in ST, as in upper-limb post stroke activations [30], [31]. *β*-CMC is more evident over parieto-occipital and parieto-central channels (e.g. CP2, CP2, PO3, Pz and PO4) during exoskeleton-assisted walking in ST. More posterior (parietal) topography of *β*-CMC suggests an increased somatosensory input during EKSO walking in ST and postural control and search for stability via proprioceptive integration [57]. Additional sensory feedback during exoskeleton use seems to be a feature and may be considered an active part of neurorehabilitation and possible gait restoration.

The final research question asks if gait-parameters, ERSP, and/or CMC improve efficiently during robot assisted gait after stroke. Our findings suggest that an inefficient improvement occurs in gait-parameters, notably swing-time, during exoskeleton use in stroke survivors. This improvement is likely not neurally-driven because it is not in unison (i.e., non-correlated and non-aligned) with any increment on neural desynchronization or CMC during a particular gait-event.

During exoskeleton use in stroke survivors, we can infer that the gait-parameter changes we observed may be merely mechanical and driven by the device itself rather than a more neural drive. The somatosensory feedback provided by the exoskeleton device is not efficiently propagated at high-level neural structures. This effect is supported by the correlations reported in Figure 3 and Table IV that describe the absence of association between the gait-parameter changes and increments of *θ*-desynchronization and *β*-CMC.

While some correlations were observed with TA and RF muscle activity, these were only evident in the unaffected limb in stroke participants. The exoskeleton provided an adequate synergy-support for maintaining a stable stance, and a swing-time reduction during gait, however, the corresponding gait-parameter improvement is unlikely considered neurally-driven [19], [56], [57] Other types of rehabilitation such as repetitive isometric movements and/or Functional Electrical Stimulation (FES) may contribute to a better sensory feedback propagation when used in RAGT [5], [16], [34], [58], [57]. Additional research is required to test multiple exoskeleton modes of assistance and an increment on movement repetitions is necessary to confirm if the minimal brain-muscle connection observed in this study in stroke can potentially help restore healthy-gait in stroke participants. This study did not address this question and currently there is not enough conclusive evidence to suggest potential for healthy-gait restoration after stroke arising from extended Exoskeleton walking therapy.

## V. Conclusion

This study measures ERSP and CMC during gait with and without an exoskeleton in healthy individuals and post-stroke survivors. While clear ERSP patterns and *β*-CMC related to the gait cycle are evident in overground and exoskeleton walking in healthy individuals, results from our stroke participants remain inconclusive as to whether walking in an exoskeleton would result in brain oscillatory dynamics as observed in healthy individuals.

While exoskeleton use in stroke altered parameters of gait that included a reduction in swing time, ERSP and CMC measures suggest these changes are not a result of improved neural efficiency. The changes on ERSP and CMC are not aligned with gait-parameters changes. The high *θ*-desynchronization and *β*-CMC associated with the use of exoskeleton in stroke participants were not correlated with the swing time reduction. Healthy participants show a different neural pattern during free overground and exoskeleton walking. The increments of gait-performance and the increments of neural desynchronization and CMC measures suggest there is no immediate modulation of neural measures during a RAGT session.

This study was limited to exploring the immediate effects of exoskeleton gait on ERSP and CMC variables during an exoskeleton walking session and was not designed to examine the effects of multiple training sessions with more movement repetitions.

## Data Availability

Data is under regulations and accessibility conditions established by University College Dublin (UCD), School of Public Health, Physiotherapy and Sports Science at UCD, and professor Olive Lennon, PhD.

## VI. Acknowledgements

This study was funded by the European Union Horizon 2020 Research and Innovation programme under the Marie Sklodowska-Curie grant agreement no.778043 called Progait, the Science Foundation Ireland, Frontiers for the Future award SFI Award 19/FFP/6747, and the Italian Ministry for foreign Affairs and International Cooperation grant No.PGR-01045 (SoftAct project). No clinical trial ID has been associated to this study in clinicaltrial.gov. This study is not planning to evaluate any treatment effect related to the Ekso device walking-assistance in stroke patients, but the variability and interaction of the outcome measures evaluated here. Clinically-related IDs associated with this study can be only referred as the grant/awards numbers reported above. We are grateful for access to the Tier 2 High Performance Computing resources provided by the Northern Ireland High Performance Computing (NI-HPC) facility funded by the UK Engineering and Physical Sciences Research Council (EPSRC), Grant Nos. EP/T022175/ and EP/W03204X/1.

## References

[1] N. S. Ward, “Restoring brain function after stroke—bridging the gap between animals and humans,” Nature Reviews Neurology, vol 13, no. 4, pp. 244–255, 2017.

[2] S. Mendis, S. Davis, and B. Norrving, “Organizational update: the world health organization global status report on noncommunicable diseases 2014; one more landmark step in the combat against stroke and vascular disease,” Stroke, vol 46, no. 5, pp. e121–e122, 2015.

[3] Y. He, D. Eguren, J. M. Azorín, R. G. Grossman, T. P. Luu, and J. L. Contreras-Vidal, “Brain–machine interfaces for controlling lower-limb powered robotic systems,” Journal of neural engineering, vol 15, no. 2, p. 021004, 2018.

[4] B. Norrving, J. Barrick, A. Davalos, M. Dichgans, C. Cordonnier, A. Guekht, K. Kutluk, R. Mikulik, J. Wardlaw, E. Richard et al., “Action plan for stroke in europe 2018–2030,” European Stroke Journal, vol 3, no. 4, pp. 309–336, 2018.

[5] O. Lennon, M. Tonellato, A. Del Felice, R. Di Marco, C. Fingleton, A. Korik, E. Guanziroli, F. Molteni, C. Guger, R. Otner et al., “A systematic review establishing the current state-of-the-art, the limitations, and the desired checklist in studies of direct neural interfacing with robotic gait devices in stroke rehabilitation,” Frontiers in Neuroscience, vol 14, p. 578, 2020.

[6] M. A. Cervera, S. R. Soekadar, J. Ushiba, J. d. R. Millán, M. Liu, N. Birbaumer, and G. Garipelli, “Brain-computer interfaces for poststroke motor rehabilitation: a meta-analysis,” Annals of clinical and translational neurology, vol 5, no. 5, pp. 651–663, 2018.

[7] Y. He, K. Nathan, A. Venkatakrishnan, R. Rovekamp, C. Beck, R. Ozdemir, G. E. Francisco, and J. L. Contreras-Vidal, “An integrated neuro-robotic interface for stroke rehabilitation using the nasa x1 powered lower limb exoskeleton,” in 2014 36th Annual International Conference of the IEEE Engineering in Medicine and Biology Society. IEEE, 2014, pp. 3985–3988.

[8] M. Dee, O. Lennon, and C. O’Sullivan, “A systematic review of physical rehabilitation interventions for stroke in low and lower-middle income countries,” Disability and rehabilitation, vol 42, no. 4, pp. 473–501, 2020.

[9] J. Bernhardt, K. S. Hayward, G. Kwakkel, N. S. Ward, S. L. Wolf, K. Borschmann, J. W. Krakauer, L. A. Boyd, S. T. Carmichael, D. Corbett et al., “Agreed definitions and a shared vision for new standards in stroke recovery research: the stroke recovery and rehabilitation roundtable taskforce,” International Journal of Stroke, vol 12, no. 5, pp. 444–450, 2017.

[10] J. Mehrholz, S. Thomas, C. Werner, J. Kugler, M. Pohl, and B. Elsner, “Electromechanical-assisted training for walking after stroke: a major update of the evidence,” Stroke, vol 48, no. 8, pp. e188–e189, 2017.

[11] R. Di Marco, M. Rubega, O. Lennon, E. Formaggio, N. Sutaj, G. Dazzi, C. Venturin, I. Bonini, R. Ortner, H. A. Cerrel Bazo et al., “Experimental protocol to assess neuromuscular plasticity induced by an exoskeleton training session,” Methods and Protocols, vol 4, no. 3, p. 48, 2021.

[12] R. S. Calabro`, A. Naro, M. Russo, P. Bramanti, L. Carioti, T. Balletta, A. Buda, A. Manuli, S. Filoni, and A. Bramanti, “Shaping neuroplasticity by using powered exoskeletons in patients with stroke: a randomized clinical trial,” Journal of neuroengineering and rehabilitation, vol 15, no. 1, pp. 1–16, 2018.

[13] S. Srivastava, P.-C. Kao, S. H. Kim, P. Stegall, D. Zanotto, J. S. Higginson, S. K. Agrawal, and J. P. Scholz, “Assist-as-needed robot-aided gait training improves walking function in individuals following stroke,” IEEE Transactions on Neural Systems and Rehabilitation Engineering, vol 23, no. 6, pp. 956–963, 2014.

[14] J. L. Contreras-Vidal, M. Bortole, F. Zhu, K. Nathan, A. Venkatakrishnan, G. E. Francisco, R. Soto, and J. L. Pons, “Neural decoding of robot-assisted gait during rehabilitation after stroke,” American journal of physical medicine & rehabilitation, vol 97, no. 8, pp. 541–550, 2018.

[15] J. T. Gwin, K. Gramann, S. Makeig, and D. P. Ferris, “Electrocortical activity is coupled to gait cycle phase during treadmill walking,” Neuroimage, vol 54, no. 2, pp. 1289–1296, 2011.

[16] J. Wagner, S. Makeig, M. Gola, C. Neuper, and G. Müller-Putz, “Distinct β band oscillatory networks subserving motor and cognitive control during gait adaptation,” Journal of Neuroscience, vol 36, no. 7, pp. 2212–2226, 2016.

[17] M. Severens, B. Nienhuis, P. Desain, and J. Duysens, “Feasibility of measuring event related desynchronization with electroencephalography during walking,” in 2012 Annual International Conference of the IEEE Engineering in Medicine and Biology Society. IEEE, 2012, pp. 2764–2767.

[18] A. T. Winslow, J. Brantley, F. Zhu, J. L. C. Vidal, and H. Huang, “Corticomuscular coherence variation throughout the gait cycle during overground walking and ramp ascent: a preliminary investigation,” in 2016 38th Annual International Conference of the IEEE Engineering in Medicine and Biology Society (EMBC). IEEE, 2016, pp. 4634–4637.

[19] M. Rubega, R. Di Marco, M. Zampini, E. Formaggio, E. Menegatti, P. Bonato, S. Masiero, and A. Del Felice, “Muscular and cortical activation during dynamic and static balance in the elderly: A scoping review,” Aging Brain, vol 1, p. 100013, 2021.

[20] Z. Guo, Q. Qian, K. Wong, H. Zhu, Y. Huang, X. Hu, and Y. Zheng, “Altered corticomuscular coherence (cmcoh) pattern in the upper limb during finger movements after stroke,” Frontiers in Neurology, vol 11, p. 410, 2020.

[21] J. Wagner, T. Solis-Escalante, P. Grieshofer, C. Neuper, G. Müller-Putz, and R. Scherer, “Level of participation in robotic-assisted treadmill walking modulates midline sensorimotor eeg rhythms in able-bodied subjects,” Neuroimage, vol 63, no. 3, pp. 1203–1211, 2012.

[22] E. García-Cossio, M. Severens, B. Nienhuis, J. Duysens, P. Desain, N. Keijsers, and J. Farquhar, “Decoding sensorimotor rhythms during robotic-assisted treadmill walking for brain computer interface (bci) applications,” PloS one, vol 10, no. 12, p. e0137910, 2015.

[23] E. Formaggio, S. Masiero, A. Bosco, F. Izzi, F. Piccione, and A. Del Felice, “Quantitative eeg evaluation during robot-assisted foot movement,” IEEE Transactions on Neural Systems and Rehabilitation Engineering, vol 25, no. 9, pp. 1633–1640, 2016.

[24] N. A. Bhagat, N. Yozbatiran, J. L. Sullivan, R. Paranjape, C. Losey, Z. Hernandez, Z. Keser, R. Grossman, G. E. Francisco, M. K. O’Malley et al., “Neural activity modulations and motor recovery following brain-exoskeleton interface mediated stroke rehabilitation,” NeuroImage: Clinical, vol 28, p. 102502, 2020.

[25] K. Knaepen, A. Mierau, E. Swinnen, H. Fernandez Tellez, M. Michielsen, E. Kerckhofs, D. Lefeber, and R. Meeusen, “Human-robot interaction: does robotic guidance force affect gait-related brain dynamics during robot-assisted treadmill walking?” PloS one, vol 10, no. 10, p. e0140626, 2015.

[26] M. Steriade, P. Gloor, R. R. Llinas, F. L. Da Silva, and M.-M. Mesulam, “Basic mechanisms of cerebral rhythmic activities,” Electroencephalography and clinical neurophysiology, vol 76, no. 6, pp. 481–508, 1990.

[27] A. De Luca, A. Bellitto, S. Mandraccia, G. Marchesi, L. Pellegrino, M. Coscia, C. Leoncini, L. Rossi, S. Gamba, A. Massone et al., “Exoskeleton for gait rehabilitation: effects of assistance, mechanical structure, and walking aids on muscle activations,” Applied Sciences, vol 9, no. 14, p. 2868, 2019.

[28] P. Vinoj, S. Jacob, V. G. Menon, S. Rajesh, and M. R. Khosravi, “Brain-controlled adaptive lower limb exoskeleton for rehabilitation of poststroke paralyzed,” IEEE Access, vol 7, pp. 132 628–132 648, 2019.

[29] J. T. Gwin and D. P. Ferris, “Beta-and gamma-range human lower limb corticomuscular coherence,” Frontiers in human neuroscience, vol 6, p. 258, 2012.

[30] H. E. Rossiter, C. Eaves, E. Davis, M.-H. Boudrias, C.-h. Park, S. Farmer, G. Barnes, V. Litvak, and N. S. Ward, “Changes in the location of cortico-muscular coherence following stroke,” NeuroImage: Clinical, vol 2, pp. 50–55, 2013.

[31] K. von Carlowitz-Ghori, Z. Bayraktaroglu, F. U. Hohlefeld, F. Losch, G. Curio, and V. V. Nikulin, “Corticomuscular coherence in acute and chronic stroke,” Clinical Neurophysiology, vol 125, no. 6, pp. 1182–1191, 2014.

[32] S.-C. Bao, W.-C. Leung, V. C. Cheung, P. Zhou, and K.-Y. Tong, “Pathway-specific modulatory effects of neuromuscular electrical stimulation during pedaling in chronic stroke survivors,” Journal of neuro-engineering and rehabilitation, vol 16, no. 1, pp. 1–15, 2019.

[33] M. G. Maggio, A. Naro, A. Manuli, G. Maresca, T. Balletta, D. Latella, R. De Luca, and R. S. Calabro, “Effects of robotic neurorehabilitation on body representation in individuals with stroke: A preliminary study focusing on an eeg-based approach,” Brain Topography, vol 34, no. 3, pp. 348–362, 2021.

[34] M. Seeber, R. Scherer, J. Wagner, T. Solis-Escalante, and G. R. Müller-Putz, “Eeg beta suppression and low gamma modulation are different elements of human upright walking,” Frontiers in human neuroscience, vol 8, p. 485, 2014.

[35] L. Brenner, “Exploring the psychosocial impact of ekso bionics technology,” Archives of Physical Medicine and Rehabilitation, vol 97, no. 10, p. e113, 2016.

[36] J. M. M. Torres, T. Clarkson, K. M. Hauschild, C. C. Luhmann, M. D. Lerner, and G. Riccardi, “Facial emotions are accurately encoded in the neural signal of those with autism spectrum disorder: A deep learning approach,” Biological Psychiatry: Cognitive Neuroscience and Neuroimaging, 2021.

[37] J. M. Mayor-Torres, M. Ravanelli, S. E. Medina-DeVilliers, M. D. Lerner, and G. Riccardi, “Interpretable sincnet-based deep learning for emotion recognition from eeg brain activity,” in 2021 43rd Annual International Conference of the IEEE Engineering in Medicine & Biology Society (EMBC). IEEE, 2021, pp. 412–415.

[38] J. M. M. Torres, T. Clarkson, E. A. Stepanov, C. C. Luhmann, M. D. Lerner, and G. Riccardi, “Enhanced error decoding from error-related potentials using convolutional neural networks,” in 2018 40th annual international conference of the IEEE engineering in medicine and biology society (EMBC). IEEE, 2018, pp. 360–363.

[39] J. M. M. Torres, “Eeg signals classification using linear and non-linear discriminant methods,” El Hombre y la Máquina, no. 41, pp. 71–80, 2013.

[40] H. M. O’Leary, W. E. Kaufmann, K. V. Barnes, K. Rakesh, K. Kapur, D. C. Tarquinio, N. G. Cantwell, K. J. Roche, S. A. Rose, A. C. Walco et al., “Placebo-controlled crossover assessment of mecasermin for the treatment of rett syndrome,” Annals of Clinical and Translational Neurology, vol 5, no. 3, pp. 323–332, 2018.

[41] J. M. M. Torres, E. A. Stepanov, and G. Riccardi, “Eeg semantic decoding using deep neural networks,” in Rovereto Workshop on Concepts, Actions, and Objects (CAOS) in Rovereto Italy, May 2016.

[42] D. Stegeman and H. Hermens, “Standards for surface electromyography: The european project surface emg for non-invasive assessment of muscles (seniam),” Enschede: Roessingh Research and Development, vol 10, pp. 8–12, 2007.

[43] T. J. Quinn, J. Dawson, M. R. Walters, and K. R. Lees, “Reliability of the modified rankin scale: a systematic review,” Stroke, vol 40, no. 10, pp. 3393–3395, 2009.

[44] J. Mehrholz, K. Wagner, K. Rutte, D. Meiβner, and M. Pohl, “Predictive validity and responsiveness of the functional ambulation category in hemiparetic patients after stroke,” Archives of physical medicine and rehabilitation, vol 88, no. 10, pp. 1314–1319, 2007.

[45] N. Bigdely-Shamlo, T. Mullen, C. Kothe, K.-M. Su, and K. A. Robbins, “The prep pipeline: standardized preprocessing for large-scale eeg analysis,” Frontiers in neuroinformatics, vol 9, p. 16, 2015.

[46] A. Mognon, J. Jovicich, L. Bruzzone, and M. Buiatti, “Adjust: An automatic eeg artifact detector based on the joint use of spatial and temporal features,” Psychophysiology, vol 48, no. 2, pp. 229–240, 2011.

[47] A. Delorme and S. Makeig, “Eeglab: an open source toolbox for analysis of single-trial eeg dynamics including independent component analysis,” Journal of neuroscience methods, vol 134, no. 1, pp. 9–21, 2004.

[48] G. Gómez-Herrero, “Automatic artifact removal (aar) toolbox v1. 3 (release 09.12. 2007) for matlab,” Tampere University of Technology, 2007.

[49] A. S. Oliveira, B. R. Schlink, W. D. Hairston, P. König, and D. P. Ferris, “Induction and separation of motion artifacts in eeg data using a mobile phantom head device,” Journal of neural engineering, vol 13, no. 3, p. 036014, 2016.

[50] M. W. Flood, B. P. O’Callaghan, and M. M. Lowery, “Gait event detection from accelerometry using the teager–kaiser energy operator,” IEEE Transactions on Biomedical Engineering, vol 67, no. 3, pp. 658–666, 2019.

[51] B. P. O’Callaghan, E. P. Doheny, C. Goulding, E. Fortune, and M. M. Lowery, “Adaptive gait segmentation algorithm for walking bout detection using tri-axial accelerometers,” in 2020 42nd Annual International Conference of the IEEE Engineering in Medicine & Biology Society (EMBC). IEEE, 2020, pp. 4592–4595.

[52] G. Pfurtscheller, “Functional brain imaging based on erd/ers,” Vision research, vol 41, no. 10-11, pp. 1257–1260, 2001.

[53] S. Makeig, “Auditory event-related dynamics of the eeg spectrum and effects of exposure to tones,” Electroencephalography and clinical neurophysiology, vol 86, no. 4, pp. 283–293, 1993.

[54] J. Rosenberg, A. Amjad, P. Breeze, D. Brillinger, and D. Halliday, “The fourier approach to the identification of functional coupling between neuronal spike trains,” Progress in biophysics and molecular biology, vol 53, no. 1, pp. 1–31, 1989.

[55] D. Halliday, J. Rosenberg, A. Amjad, P. Breeze, B. Conway, S. Farmer et al., “A framework for the analysis of mixed time series/point process data-theory and application to the study of physiological tremor, single motor unit discharges and electromyograms,” Progress in biophysics and molecular biology, vol 64, no. 2, p. 237, 1995.

[56] F. Molteni, E. Formaggio, A. Bosco, E. Guanziroli, F. Piccione, S. Masiero, and A. Del Felice, “Brain connectivity modulation after exoskeleton-assisted gait in chronic hemiplegic stroke survivors: A pilot study,” American journal of physical medicine & rehabilitation, vol 99, no. 8, pp. 694–700, 2020.

[57] T. Solis-Escalante, J. van der Cruijsen, D. de Kam, J. van Kordelaar, V. Weerdesteyn, and A. C. Schouten, “Cortical dynamics during preparation and execution of reactive balance responses with distinct postural demands,” NeuroImage, vol 188, pp. 557–571, 2019.

[58] J. Wagner, T. Solis-Escalante, R. Scherer, C. Neuper, and G. MüllerPutz, “It’s how you get there: walking down a virtual alley activates premotor and parietal areas,” Frontiers in human neuroscience, vol 8, p. 93, 2014.

[59] H. Enders and B. M. Nigg, “Measuring human locomotor control using emg and eeg: Current knowledge, limitations and future considerations,” European journal of sport science, vol 16, no. 4, pp. 416–426, 2016.

