## supplementary material for "Robotic-Assisted Gait for lower-limb Rehabilitation: Evidence of Altered Neural Mechanisms in Stroke"

Juan Manuel Mayor-Torres, Ben O'Callaghan, Attila Korik, Alessandra Del Felice, *Member, IEEE,* Damien Coyle, *Member, IEEE,* Sean Murphy, and Olive Lennon

### SUPPLEMENTARY MATERIAL

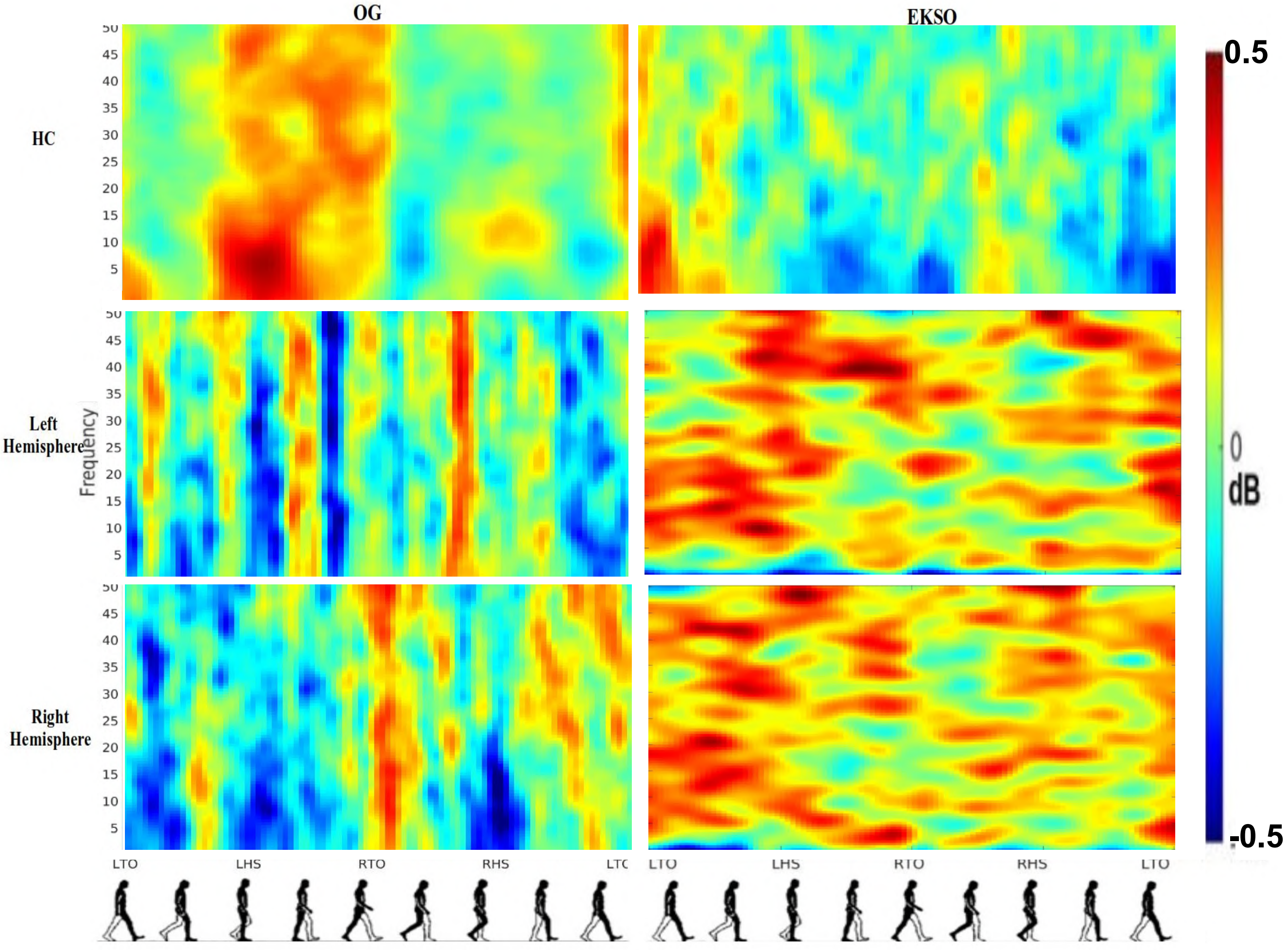


Fig S 1: Average ERSP output for the Cz channel. This Figure shows in rows the groups - healthy-controls (HC), and Stroke Patients (ST) Left Stroke side and Right Stroke side, and in columns the gait-evaluation modalities OG and Ekso. Each plot has in y-axis frequency between 0-50Hz, and in x-axis all the gait-phases of the gait-cycle. Red spots represent synchronization and darker blue spots represent desynchronization. The level of sync-desync is defined in the colorbar between [-0.5,0.5] dB and plotted based on the jet colormap as Knaepen et al. [1] suggested.

Juan Manuel Mayor-Torres is at School of Public Health, Physiotherapy and Sports Science at University College Dublin, Belfield, Dublin,**, Ben O'Callaghan is at School of Public Health, Physiotherapy and Sports Science at University College Dublin, Belfield, Dublin,. Attila Korik is at School of Computing, Engineering and Intelligent Systems at Ulster University,, Alessandra Del Felice is the head of Neurophysiology and Movement Rehabilitation group and at Padova Neuroscience Center (PNC) at University of Padova,, Damien Coyle is at School of Computing, Engineering and Intelligent Systems at Ulster University,, Sean Murphy is at Mater Misericordiae Hospital, Dublin,, Olive Lennon is at School of Public Health, Physiotherapy and Sports Science at University College Dublin, Belfield, Dublin,

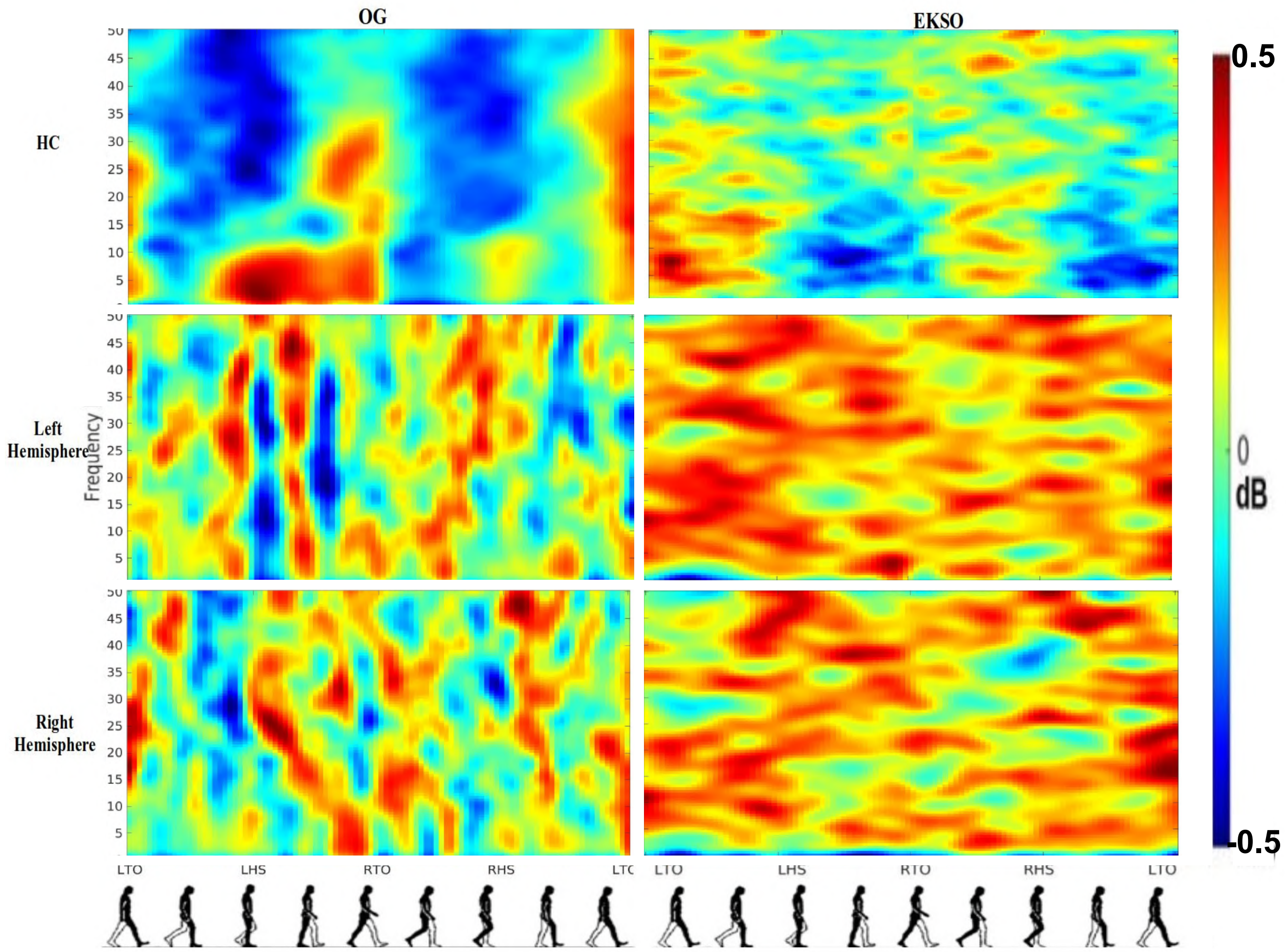


Fig S 2: Average ERSP output for the C4 channel. This Figure shows in rows the groups - healthy-controls (HC), and Stroke Patients (ST) Left Stroke side and Right Stroke side, and in columns the gait-evaluation modalities OG and Ekso. Each plot has in y-axis frequency between 0-50Hz, and in x-axis all the gait-phases of the gait-cycle. Red spots represent synchronization and darker blue spots represent desynchronization. The level of sync-desync is defined in the colorbar between [-0.5,0.5] dB and plotted based on the jet colormap as Knaepen et al. [1] suggested.

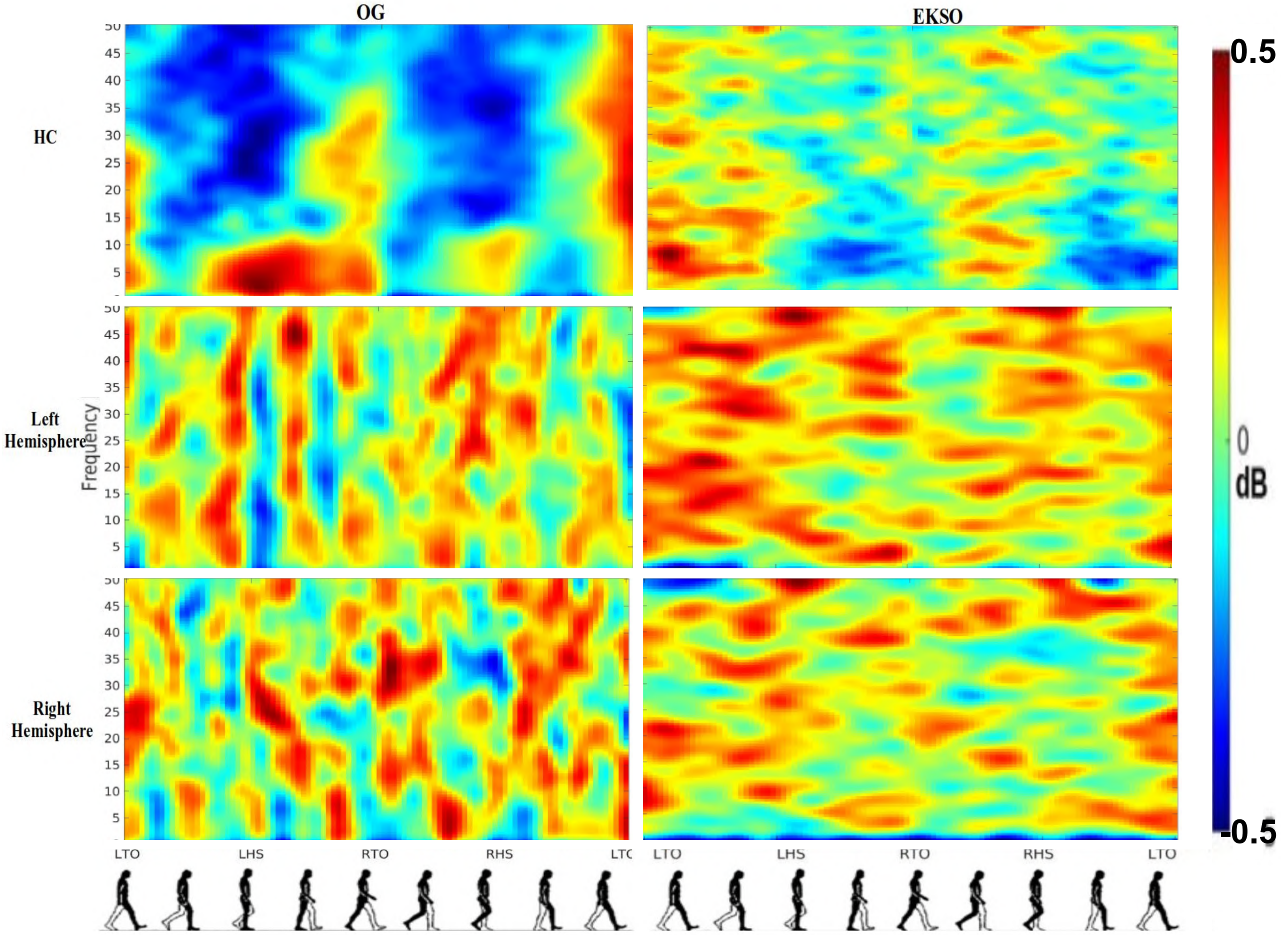


Fig S 3: Average ERSP output for the C3 channel. This Figure shows in rows the groups - healthy-controls (HC), and Stroke Patients (ST) Left Stroke side and Right Stroke side, and in columns the gait-evaluation modalities OG and Ekso. Each plot has in y-axis frequency between 0-50Hz, and in x-axis all the gait-phases of the gait-cycle. Red spots represent synchronization and darker blue spots represent desynchronization. The level of sync-desync is defined in the colorbar between [-0.5,0.5] dB and plotted based on the jet colormap as Knaepen et al. [1] suggested.

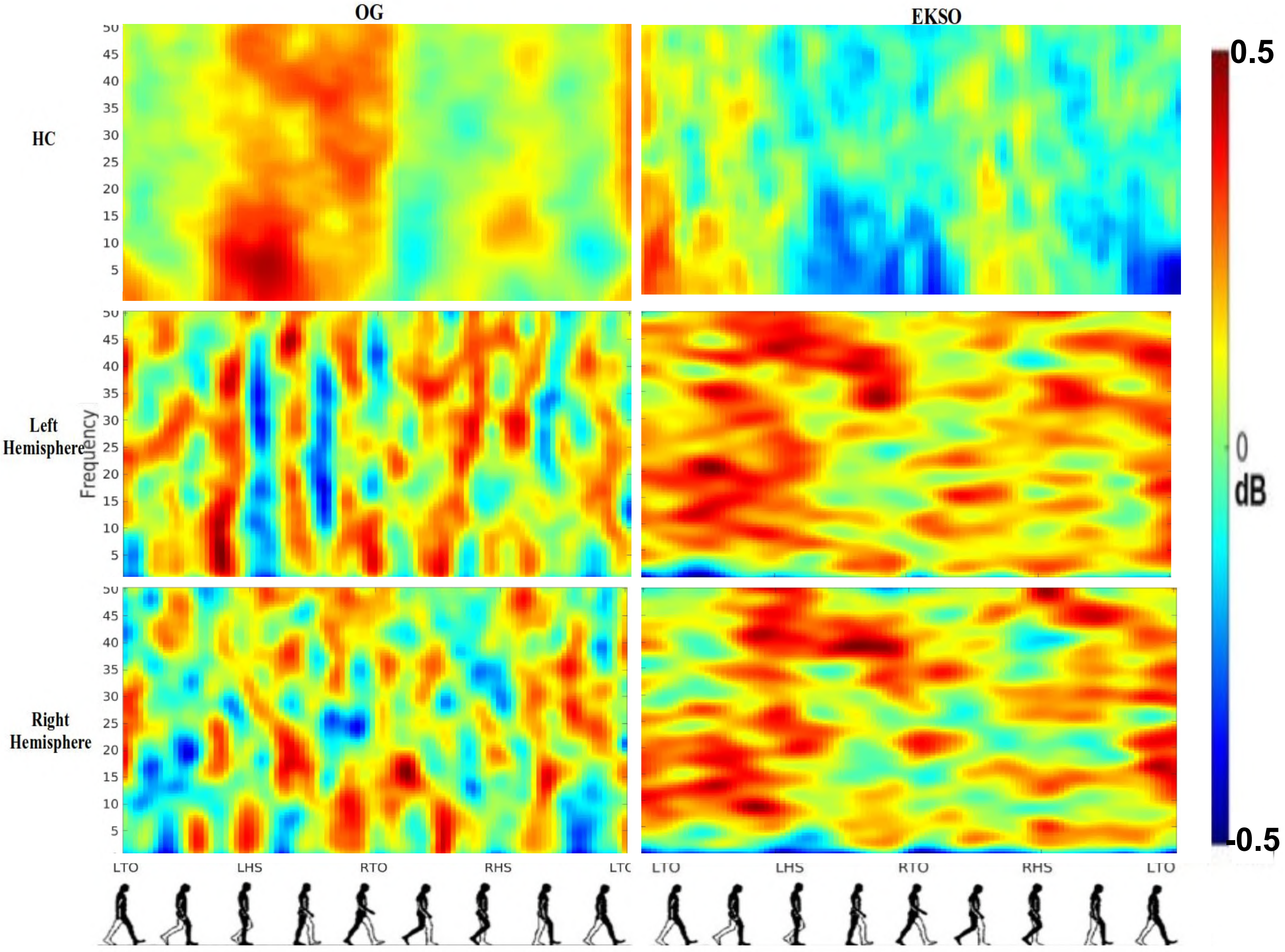


Fig S 4: Average ERSP output for the Pz channel. This Figure shows in rows the groups - healthy-controls (HC), and Stroke Patients (ST) Left Stroke side and Right Stroke side, and in columns the gait-evaluation modalities OG and Ekso. Each plot has in y-axis frequency between 0-50Hz, and in x-axis all the gait-phases of the gait-cycle. Red spots represent synchronization and darker blue spots represent desynchronization. The level of sync-desync is defined in the colorbar between [-0.5,0.5] dB and plotted based on the jet colormap as Knaepen et al. [1] suggested.

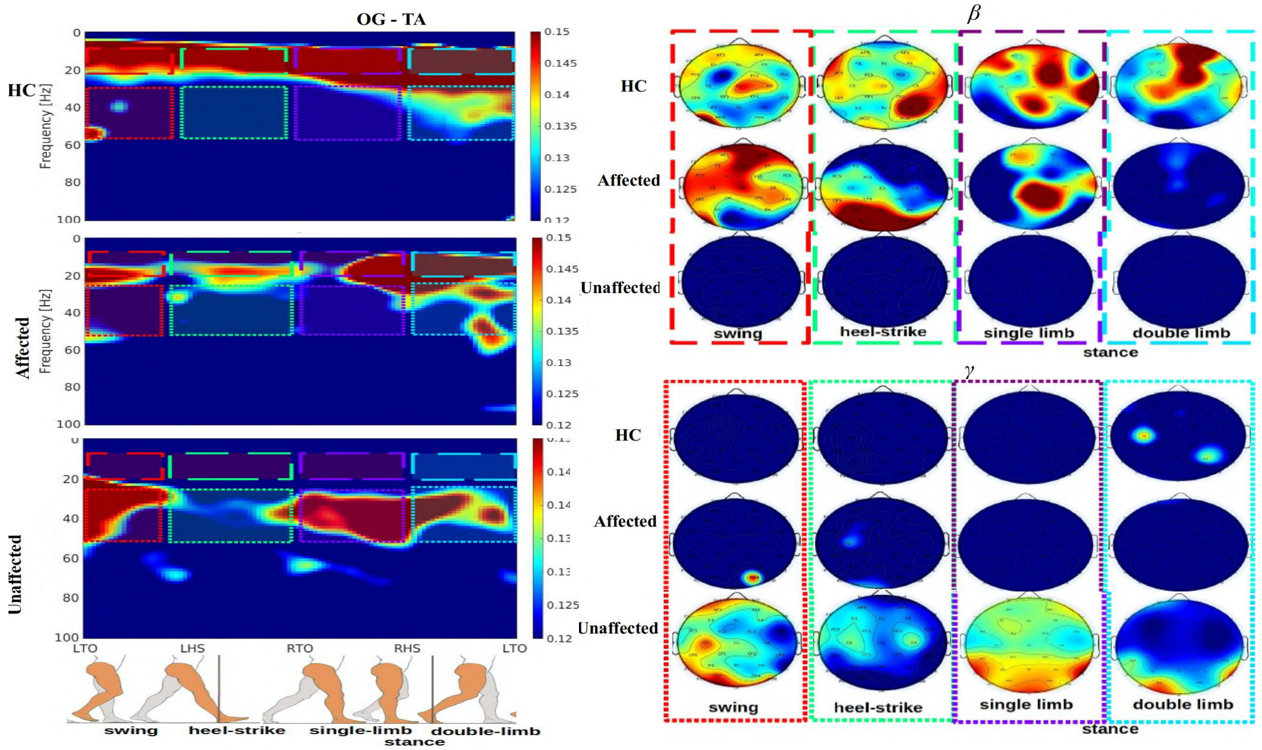


(a) Overground-Gait (OG) CMC measures - movement of the right-limb - TA left

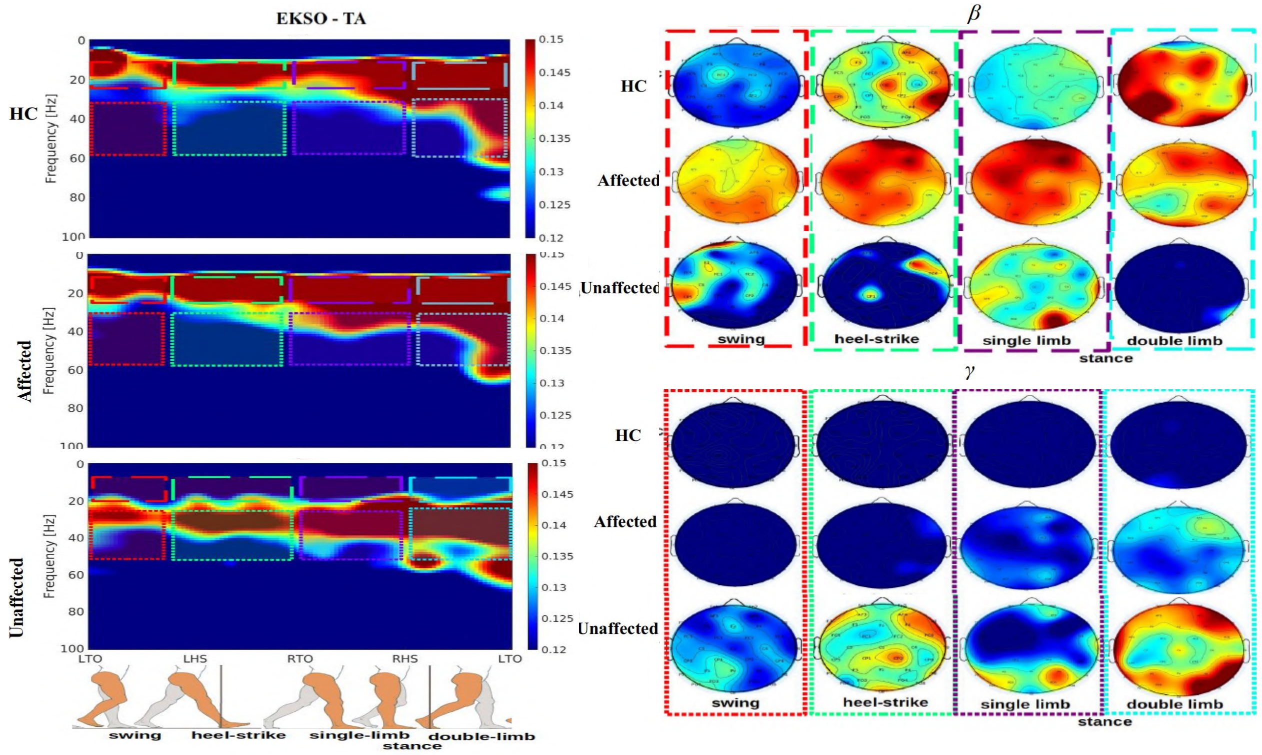


(b) Ekso CMC measures - movement of the right-limb - TA left

Fig S 5: TA left - Average CMC measures for Right Stroke Side, reporting the CMC output for healthy-controls (HC), and Stroke patients (ST) for Affected and Unaffected limb. This output is only associated withthe movement of left lower-limb. All the CMC values are plotted between [0.12,0.15] using the jet colormap and respecting the 95% of confidence interval described in we followed the methdology in [2], [3]. Figures S5a and S5b show the CMC outputs for the OG and Ekso modalities. In left plots we reported the CMC setting y-axis as frequency in Hz and x-axis as the gait-phases associated with the movement of the left limb.

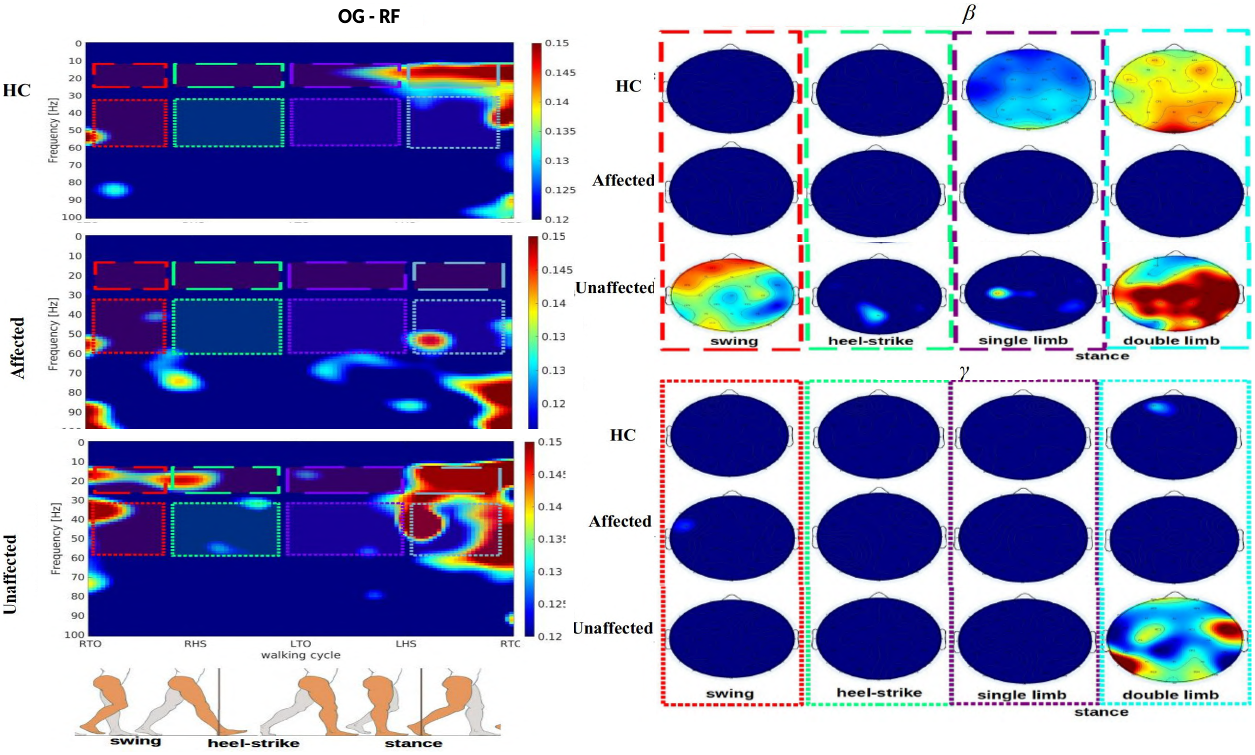


(a) Overground-Gait (OG) CMC measures - movement of the right-limb - RF right

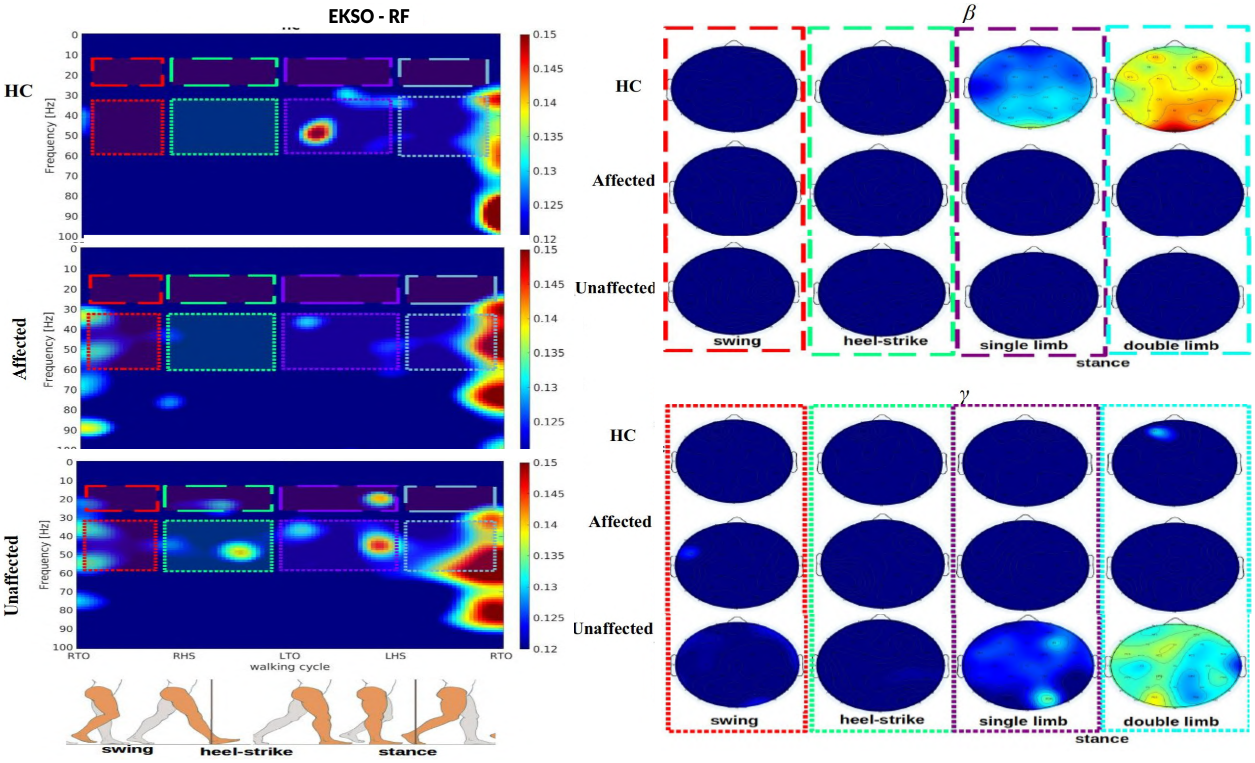


(b) Ekso CMC measures - movement of the right-limb - RF right

Fig S 6: RF right - Average CMC measures for Left Stroke Side, reporting the CMC output for healthy-controls (HC), and Stroke patients (ST) for Affected and Unaffected limb. This output is only associated with the movement of right lower-limb. All the CMC values are plotted between [0.12,0.15] using the jet colormap and respecting the 95% of confidence interval described in we followed the methdology in [2], [3]. Figures S6a and S6b show the CMC outputs for the OG and Ekso modalities. In left plots we reported the CMC setting y-axis as frequency in Hz and x-axis as the gait-phases associated with the movement of the right limb.

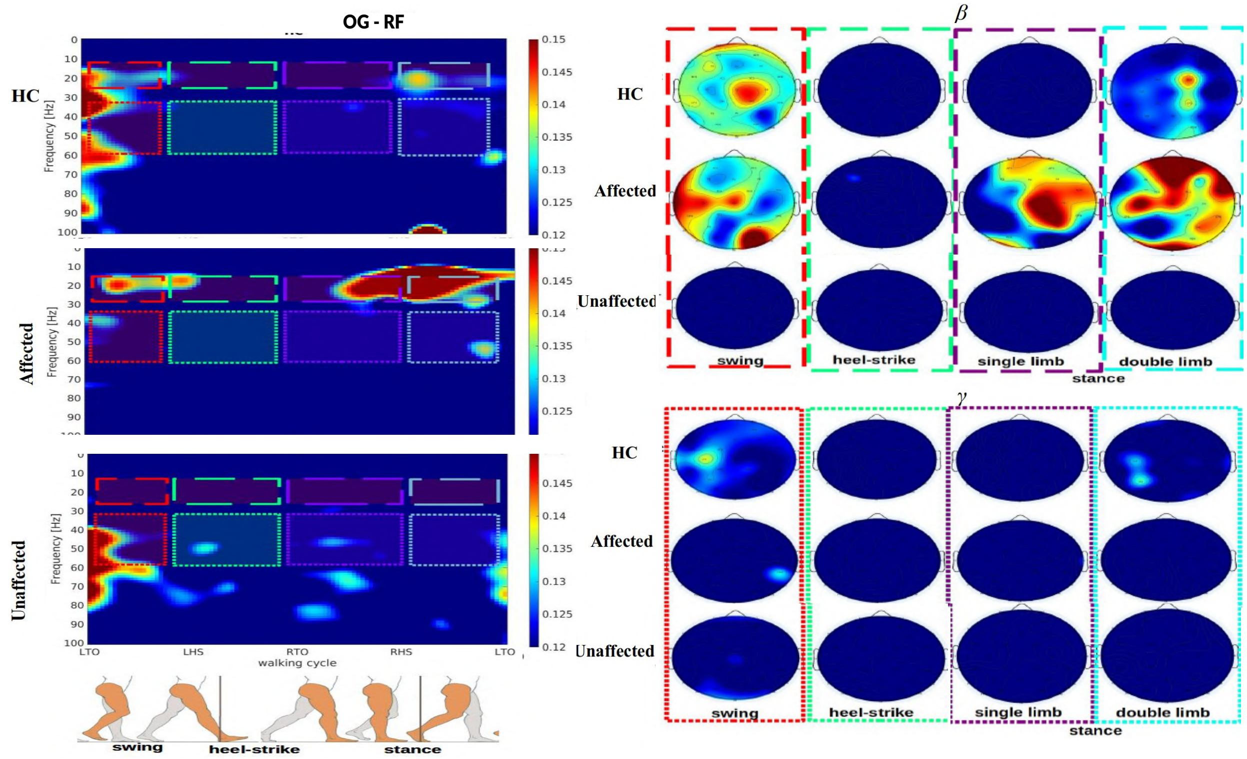


(a) Overground-Gait (OG) CMC measures - movement of the left-limb - RF left

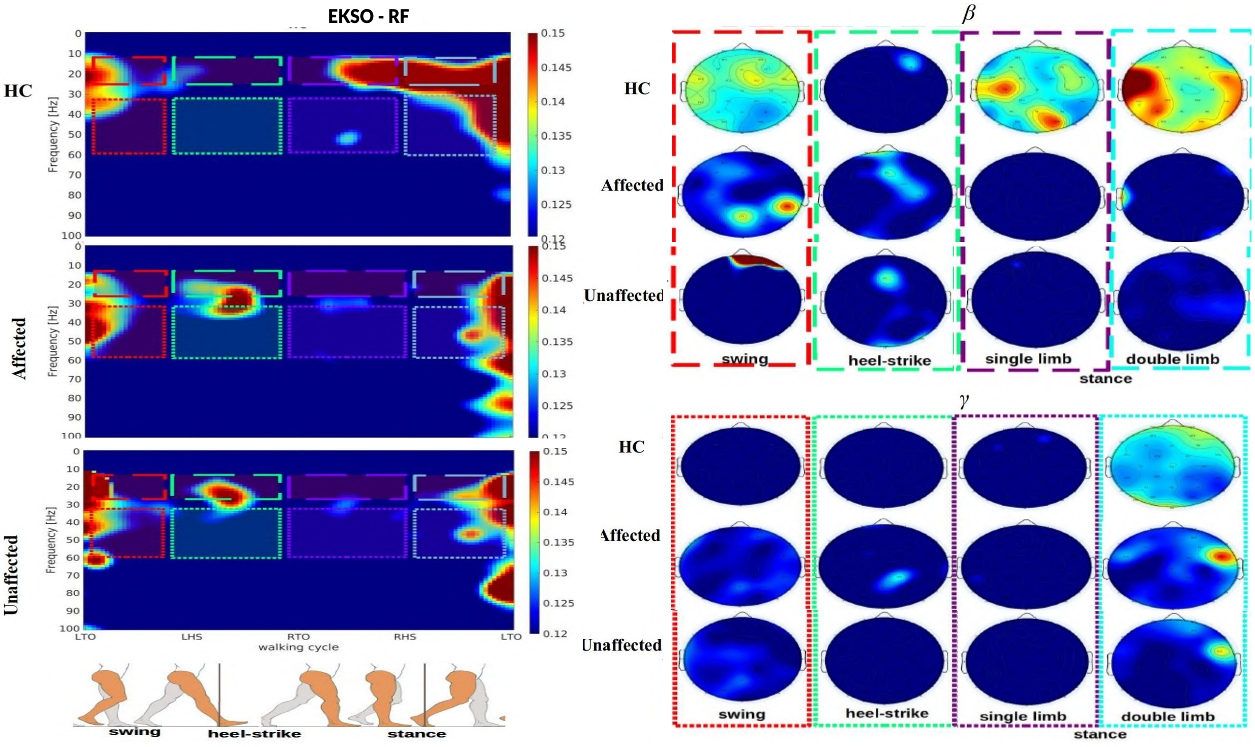


(b) Ekso CMC measures - movement of the left-limb - RF left

Fig S 7: RF left - Average CMC measures for Right Stroke Side, reporting the CMC output for healthy-controls (HC), and Stroke patients (ST) for Affected and Unaffected limb. This output is only associated with the movement of left lower-limb. All the CMC values are plotted between [0.12,0.15] using the jet colormap and respecting the 95% of confidence interval described in we followed the methdology in [2], [3]. Figures S7a and S7b show the CMC outputs for the OG and Ekso modalities. In left plots we reported the CMC setting y-axis as frequency in Hz and x-axis as the gait-phases associated with the movement of the left limb.

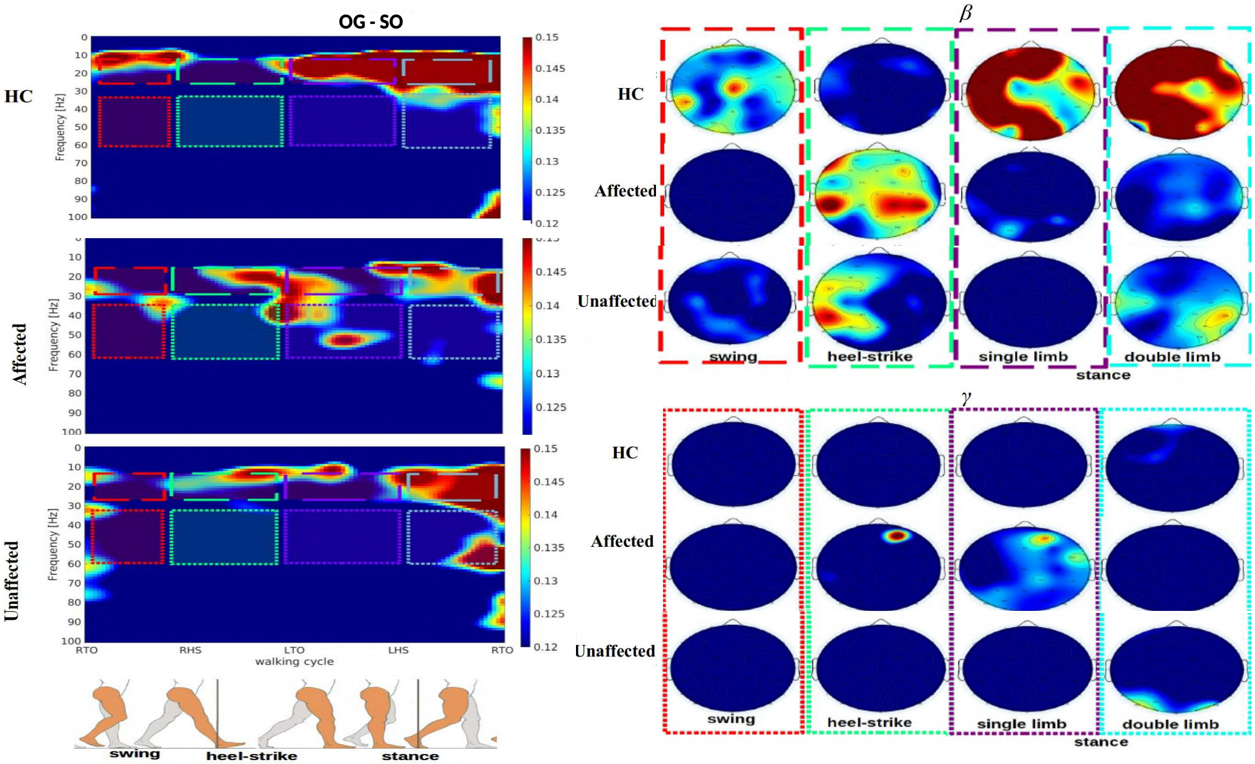


(a) Overground-Gait (OG) CMC measures - movement of the right-limb - SO right

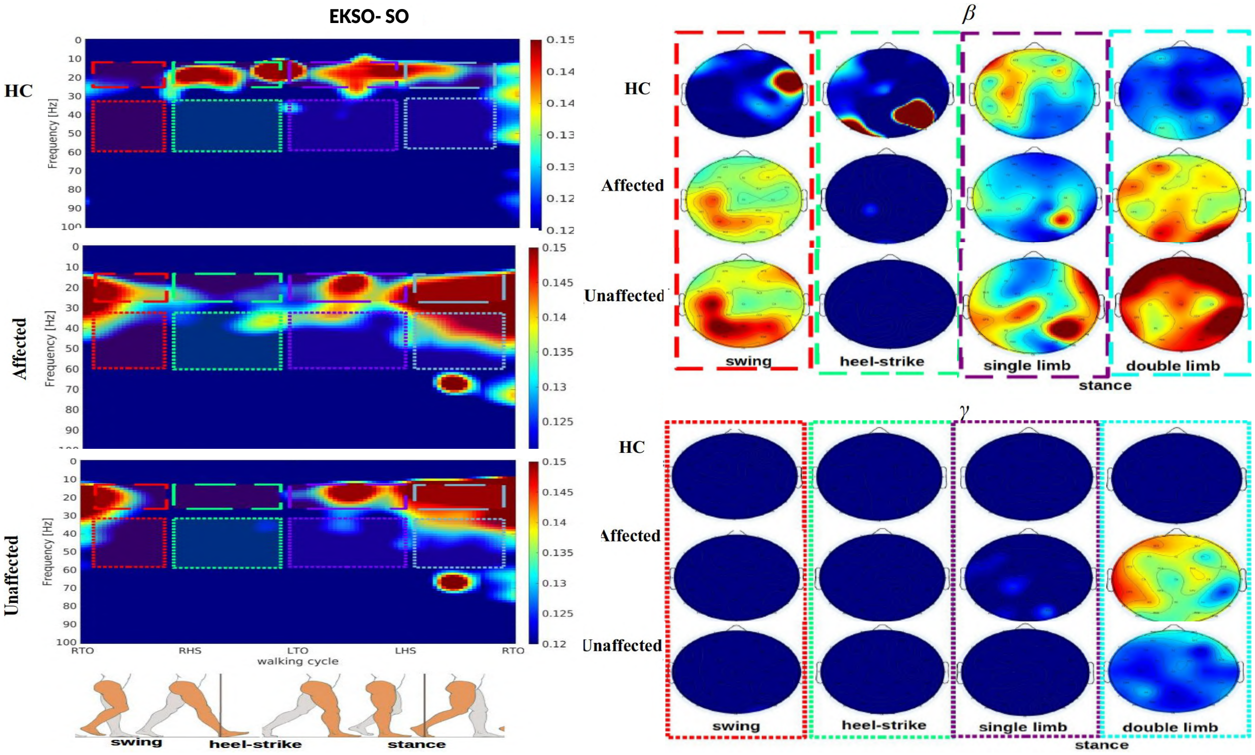


(b) Ekso CMC measures - movement of the right-limb - SO right

Fig S 8: SO right - Average CMC measures for Left Stroke Side, reporting the CMC output for healthy-controls (HC), and Stroke patients (ST) for Affected and Unaffected limb. This output is only associated with the movement of right lower-limb. All the CMC values are plotted between [0.12,0.15] using the jet colormap and respecting the 95% of confidence interval described in we followed the methdology in [2], [3]. Figures S8a and S8b show the CMC outputs for the OG and Ekso modalities. In left plots we reported the CMC setting y-axis as frequency in Hz and x-axis as the gait-phases associated with the movement of the right limb.

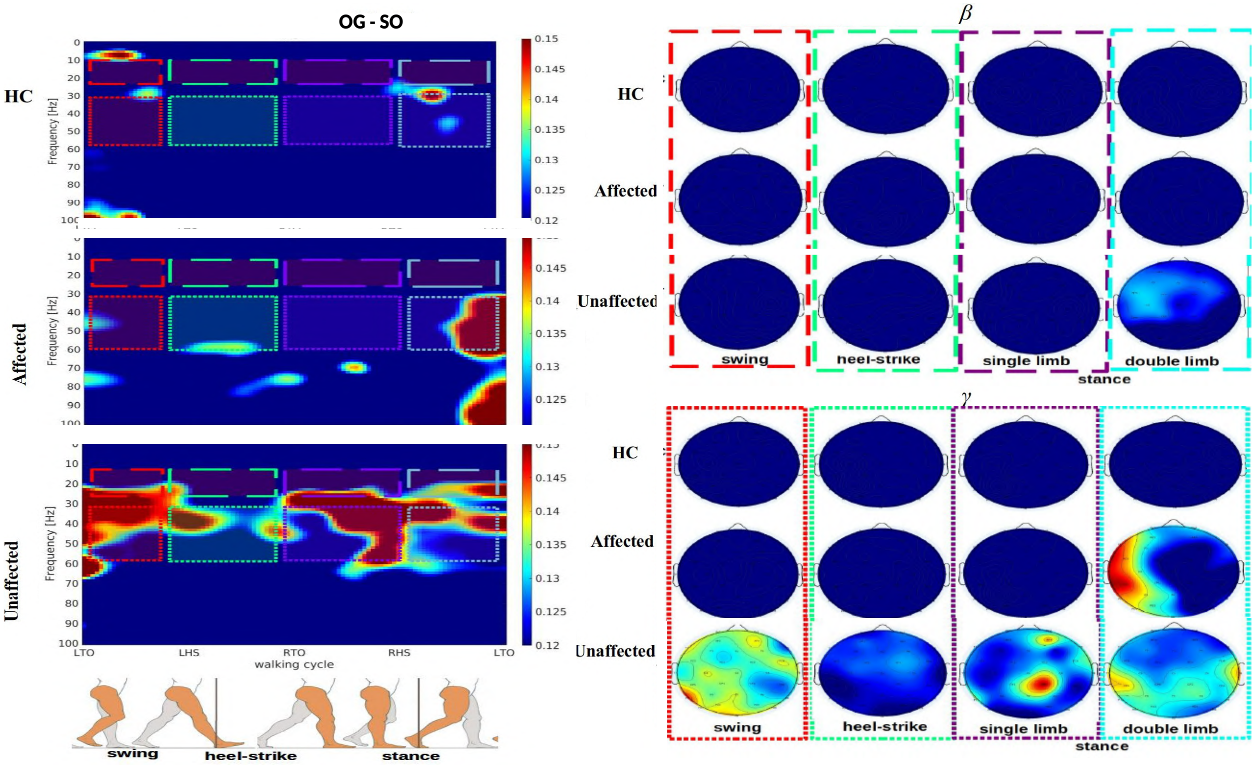


(a) Overground-Gait (OG) CMC measures - movement of the left-limb - SO left

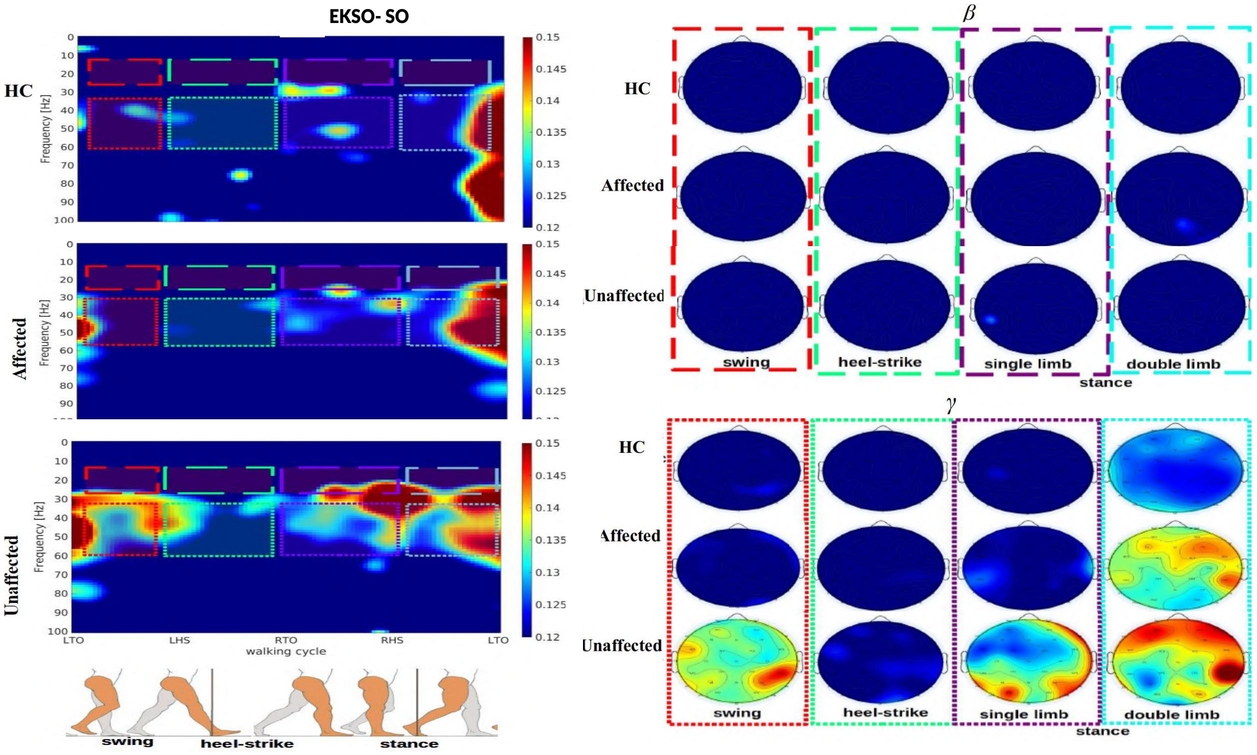


(b) Ekso CMC measures - movement of the left-limb - SO left

Fig S 9: SO left - Average CMC measures for Right Stroke Side, reporting the CMC output for healthy-controls (HC), and Stroke patients (ST) for Affected and Unaffected limb. This output is only associated with the movement of left lower-limb. All the CMC values are plotted between [0.12,0.15] using the jet colormap and respecting the 95% of confidence interval described in we followed the methdology in [2], [3]. Figures S9a and S9b show the CMC outputs for the OG and Ekso modalities. In left plots we reported the CMC setting y-axis as frequency in Hz and x-axis as the gait-phases associated with the movement of the left limb.

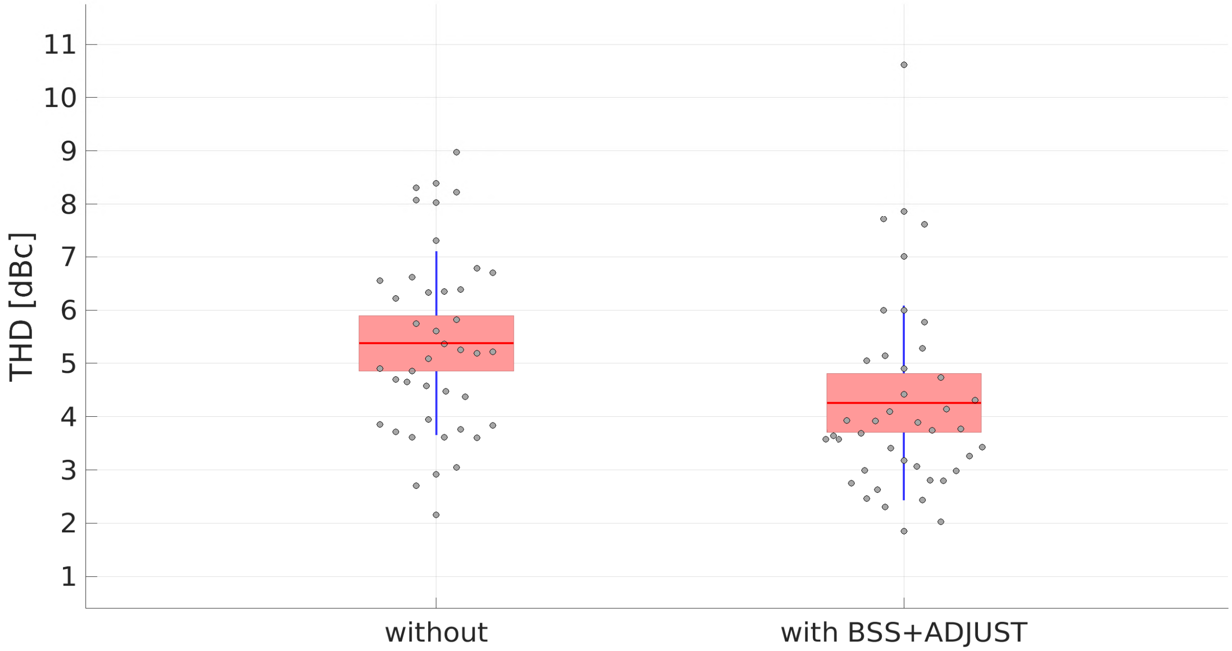


Fig S 10: Total-Harmonic Distortion (THD) analysis reporting the values in y-axis as the THD trial-values in dBc and in the x-axis we described the measures of the THD after filtering (1) without using Artifact Subspace Reconstruction (ASR) and ADJUST, and (2) using the Blind-Source Separation (BSS) method ASR and ADJUST for artifact-removal. THD difference between modalities is significant - $F(1,445)=13.35$, $p=0.0278$. The points in grey show the outlier values on each modality. For calculating the THD values in dBs we used the method described in [4].

### References

[1] K. Knaepen, A. Mierau, E. Swinnen, H. Fernandez Tellez, M. Michielsen, E. Kerckhofs, D. Lefeber, and R. Meeusen, "Human-robot interaction: does robotic guidance force affect gait-related brain dynamics during robot-assisted treadmill walking?" *PloS one*, vol. 10, no. 10, p. e0140626, 2015.

[2] H. E. Rossiter, C. Eaves, E. Davis, M.-H. Boudrias, C.-h. Park, S. Farmer, G. Barnes, V. Litvak, and N. S. Ward, "Changes in the location of cortico-muscular coherence following stroke," *NeuroImage: Clinical*, vol. 2, pp. 50–55, 2013.

[3] K. von Carlowitz-Ghori, Z. Bayraktaroglu, F. U. Hohlefeld, F. Losch, G. Curio, and V. V. Nikulin, "Corticomuscular coherence in acute and chronic stroke," *Clinical Neurophysiology*, vol. 125, no. 6, pp. 1182–1191, 2014.

[4] M. S. Diab and S. A. Mahmoud, "A 1.7 nw 24 hz variable gain elliptic low pass filter in 90-nm cmos for biosignal detection," in *2019 IEEE International Symposium on Circuits and Systems (ISCAS)*. IEEE, 2019, pp. 1–5.
